# Face Masks, Public Policies and Slowing the Spread of COVID-19: Evidence from Canada

**DOI:** 10.1101/2020.09.24.20201178

**Authors:** Alexander Karaivanov, Shih En Lu, Hitoshi Shigeoka, Cong Chen, Stephanie Pamplona

## Abstract

We estimate the impact of indoor face mask mandates and other non-pharmaceutical interventions (NPI) on COVID-19 case growth in Canada. Mask mandate introduction was staggered from mid-June to mid-August 2020 in the 34 public health regions in Ontario, Canada’s largest province by population. Using this variation, we find that mask mandates are associated with a 22 percent weekly reduction in new COVID-19 cases, relative to the trend in absence of mandate. Province-level data provide corroborating evidence. We control for mobility behaviour using Google geo-location data and for lagged case totals and case growth as information variables. Our analysis of additional survey data shows that mask mandates led to an increase of about 27 percentage points in self-reported mask wearing in public. Counterfactual policy simulations suggest that adopting a nationwide mask mandate in June could have reduced the total number of diagnosed COVID-19 cases in Canada by over 50,000 over the period July–November 2020. Jointly, our results indicate that mandating mask wearing in indoor public places can be a powerful policy tool to slow the spread of COVID-19.

**JEL codes:** I18, I12, C23

## 1 Introduction

When government policies to stem the spread of COVID-19 were introduced in early 2020, the best available supporting evidence came from research on previous epidemics, epidemiological modeling and case studies (OECD, 2020). Even when the efficacy of a given intervention in reducing viral transmission has been established, doubts regarding its usefulness may persist, because of uncertainty about adherence to the rules and other behavioural responses. For example, even though there is significant agreement in the medical literature that respiratory transmission of COVID-19 is the dominant vector (Meyerowitz et al., 2020), and many clinical studies have shown that face masks reduce the spread of COVID-19 and similar diseases (Chu et al. 2020, Prather et al. 2020, Leung et al. 2020, Greenhalgh et al. 2020), a mask mandate may not be effective in practice if it fails to increase the prevalence of mask wearing (compliance) or if it leads to increased contacts because of a false sense of security. The low economic cost of mask mandates relative to other COVID-19 containment measures has generated keen interest worldwide for studying their effectiveness.^1^ This interest has been compounded by the substantial variation in official advice regarding mask use, especially early in the pandemic, with national health authorities and the World Health Organization giving inconsistent or contradictory recommendations over time, ranging from ‘not recommended’ to ‘mandatory’.^2^ Fig. C1 in the Appendix plots self-reported mask usage in select countries (Canada, USA, Germany and Australia) and across Canadian provinces. The figure shows large differences in mask usage, both across countries and within Canada.^3^ An added challenge is to disentangle the impact of mask mandates from that of other policies, behavioural responses, or third factors (Chernozhukov et al., 2021; Mitze et al., 2020). Given the absence of large-scale randomized controlled trials on mask mandates (Howard et al., 2020), observational studies like ours are essential for informing health policy and public opinion, by formally analyzing the relationship between policy measures and the rate of propagation of COVID-19.

We estimate and quantify the impact of mask mandates and other non-pharmaceutical interventions (NPI), including regulations on businesses and gatherings, school closures, travel and self-isolation, and long-term care homes, on the growth of new COVID-19 cases in Canada. The Canadian data have the advantage of allowing two complementary approaches to address our objective. First, we estimate the effect of mandates by exploiting within-province variation in the timing of indoor face mask mandates in the 34 public health regions (Public Health Units^4^ or PHUs) in Ontario, Canada’s most populous province with roughly 40% (15 mln) of the country’s population (Statistics Canada, 2020). The advantage of this approach is that it uses variation over a relatively small geographic scale (PHU), holding all province-level policies and factors constant. The adoption of indoor face mask mandates in the 34 public health regions was staggered over approximately two months (June to August 2020), creating sufficient intertemporal variation.

Second, we evaluate the impact of NPIs in Canada as a whole, using the variation in the timing of mask mandates and other policies in the country’s ten provinces. We construct time series for the intensity and timing of COVID-relevant policy measures from official public health orders and government announcements. By studying inter-provincial variation in the policy measures, we can analyze the impact of not only mask mandates, but also other NPIs, for which there is little or no variation across Ontario’s PHUs. In addition, our data include the initial ‘closing’ period (March–April), the gradual ‘re-opening’ period (May–August), and the Fall period of new restrictions, providing variation from both the imposition and the relaxation of policies.

Our panel-data estimation strategy broadly follows the approach of Chernozhukov, Kasahara and Schrimpf (2021), which we modify and adapt to the Canadian context. We account for observed behavioural changes and trends (using Google Community Mobility Reports geolocation data as proxy), as well as for lagged outcome responses to policy and behavioural changes. Our empirical approach also allows past epidemiological outcomes to impact current outcomes, either as information variables affecting unmeasured behaviour and policy, and/or directly, as in the SIR epidemiological model (Kermack and McKendrick, 1927).

We find that, after two weeks from implementation, mask mandates are associated, on average, with a reduction of nearly 25 log points in the weekly case growth rate in Ontario, which can be interpreted as a 22% weekly reduction in new diagnosed COVID-19 cases relative to the trend in absence of mask mandate. We find corroborating evidence in the province-level analysis: a 20% weekly reduction in cases relative to the no-mandate trend in our baseline empirical specification. Furthermore, using additional survey data (Jones et al., 2021), we show that mask mandates increased self-reported mask usage in Canada by about 27 percentage points on average after implementation, confirming that the mandates had a significant impact on masking behaviour. Jointly, our results suggest that mandating mask wearing in indoor public places can be a powerful policy tool to slow the spread of COVID-19.

Counterfactual policy simulations using our empirical estimates further suggest that mandating indoor masks Canada-wide in mid-June could have reduced new COVID-19 cases in the country by more than 50,000 cases in total over the period July–November 2020 relative to the actually observed numbers.

In addition, we find that the most stringent policy restrictions on businesses and gatherings observed in the provincial data are associated with a 44% weekly decrease of new cases, relative to the trend in absence of restrictions. The business/gathering estimates are, however, noisier than our estimates for mask mandates and do not retain statistical significance in all specifications. Travel and long-term care restrictions are associated with a sharp decrease in weekly case growth in the initial closing period (March–April). These results suggest that relaxed restrictions and the associated increase in business and workplace activity and gatherings (including retail, restaurants and bars) can offset, in whole or in part, the estimated impact of mask mandates on COVID-19 case growth. We also find statistically significantly negative association between current case growth and information (log of past weekly cases), which is consistent with a behavioural feedback effect, e.g., people changing their contact rate based on observed case totals.

Our paper relates most closely to Chernozhukov et al. (2021), Lyu and Wehby (2020) and Mitze et al. (2020), which are recent studies on mask mandates using observational data, but adds to them in several ways.^5^ First, we use regional variation within the same jurisdiction (similar to Mitze et al., but with a larger sample of treated regions that obviates the need for synthetic controls). This mitigates concerns about omitted variables, e.g., provincial or other factors. Second, we additionally use variation over a different geographic and administrative level, across provinces (similar to Chernozhukov et al. and Lyu and Wehby for U.S. states), and obtain very similar results as in our main analysis. Third, we estimate the increase in self-reported mask usage following mask mandates, which can help in understanding and predicting how the effect of mask mandates on COVID-19 spread may differ in other contexts. Our finding that mask mandates led to a significant increase in mask usage corroborates the large estimated mask mandate effect on COVID-19 case growth. Fourth, an important difference between our paper and Chernozhukov et al. (2021), possibly explaining our larger estimates for the effect of mask mandates, is that we evaluate *universal* mandatory indoor mask wearing for the public, instead of mandatory mask wearing for *employees only*. While other factors such as differences in mask-wear compliance between Canada and the U.S. may also contribute to the different estimates of the policy impact, our results suggest that broader in scope mask policies can be more effective in reducing the spread of COVID-19.^6^

In the medical literature, Meyerowitz et al. (2020), a comprehensive review on COVID-19 transmission, conclude that there is strong evidence, from case and cluster reports, that respiratory transmission is dominant, with proximity and ventilation being the key determinants of transmission risk, as opposed to direct contact or fomite transmission. Numerous other studies, e.g., Prather et al. (2020), Howard et al. (2020), Greenhalgh et al. (2020), Leung et al. (2020), among others, argue that face masks can reduce the spread of COVID-19. Our paper also complements recent work on COVID-19 non-pharmaceutical interventions in various countries, e.g., Hsiang et al. (2020), Dergiades et al. (2020), Abouk and Heydari (2020) and in Canada, Mohammed et al. (2020), Yuksel et al. (2020), Armstrong et al. (2020), Stevens et al. (2021).

## 2 Data

We use three main data sources, respectively for epidemiological variables, NPIs and mask mandates, and behavioural responses. The analyzed time period is from the start of detected community transmission in Canada in March 2020 to the end of November 2020.

We located and accessed the original official data sources to compile a complete set of COVID-19 case, death, and test counts in all ten Canadian provinces over time.^7^ In addition, we collected data on cases and weekly tests for each of the 34 public health units (PHUs) in Ontario. A detailed description is provided in the data files shared at the project’s Github repository.

Implementation dates of NPIs and other public policies were collected from the official government websites, announcements, public health orders, and staged re-opening plans. In the national data, the raw policy measures data contain the dates or enactment and relaxation (if applicable) of 17 policy indicators including: mandatory mask wear; closure and re-opening of retail and non-essential businesses, restaurants, recreation facilities, and places of worship; school closures; limits on events and gatherings; international and domestic travel restrictions and self-isolation requirements; restrictions on visits and staff movement in long-term care homes. All policy indicators are defined in Table D1 in the Appendix.^8^ Since many of these indicators are highly correlated with each other, we combine them into five policy aggregates in the empirical analysis (see Table B18 and Section 3.2). In the Ontario PHU data, the implementation dates of mask mandates and the intensity of restrictions for businesses and gatherings vary across PHUs.^9^

Regarding behavioural responses, we use the Google COVID-19 Community Mobility Reports, which summarize daily cellphone geo-location data by region as indices calculated relative to the median value for the same day of the week in the five-week baseline period January 3 to February 6, 2020.^10^ In Ontario, these location data are available for each of the 51 first-level administrative divisions (counties, regional municipalities, single-tier municipalities and districts).^11^

## 3 Empirical method

We follow the approach of Chernozhukov et al. (2021), but modify and adapt it to the Canadian context. The empirical strategy uses the panel structure of the outcome, policy and behavioural proxy variables, and includes lags of outcomes, as information or following the causal paths suggested by the epidemiological SIR model (Kermack and McKendrick, 1927). Specifically, we estimate the effect of policy interventions on COVID-19 outcomes while controlling for information and behaviour. In contrast to Chernozhukov et al. (2021) and Hsiang et al. (2020), who study variation in NPIs across U.S. states or across countries, our identification strategy exploits policy variation at the sub-provincial level (Ontario’s PHUs) in addition to cross-province variation, and our data captures both the closing down and re-opening stages of the epidemic.

### 3.1 Estimation strategy

The data used in the empirical analysis are summarized below; Section 3.2 describes the variables in detail. Everywhere *i* denotes health region (PHU) for the Ontario analysis or province for the national analysis, and *t* denotes time measured in days (date).

1. Outcomes, *Y*_*it*_ – the growth rate of weekly cases or deaths.
2. Policy/NPIs, *P*_*it*_ – for the Ontario analysis, four policy aggregates by PHU and date; for the national analysis, five policy aggregates by province and date.
3. Behavioural responses, *B*_*it*_ – proxied by Google mobility data capturing changes in people’s geo-location relative to a baseline period in January-February 2020.
4. Information, *I*_*it*_ – lagged outcomes, i.e., level or growth rate of cases (or deaths). We also consider a specification that includes past cases (deaths) and case (deaths) growth at the provincial or national level as additional information variables.
5. Controls – PHU or province fixed effects, week fixed effects, the growth rate of weekly new COVID-19 tests and other possible confounders (e.g., news or weather).

To assess and disentangle the impact of mask mandates and other NPIs and behavioural responses on COVID-19 outcomes, we estimate:

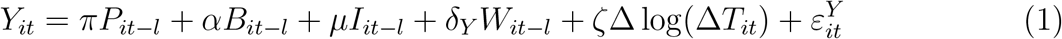

where *l* denotes time lag in days, Δ log(Δ*T*_*it*_) is the weekly growth rate of tests (defined below), and *W*_*it*−*l*_ are confounders, including fixed effects. Equation (1) models the relationship between COVID-19 outcomes, *Y*_*it*_ and lagged policy measures, *P*_*it*−*l*_, lagged behaviour, *B*_*it*−*l*_, and information (lagged outcomes), *I*_*it*−*l*_. For case growth as the outcome variable, we use a 14-day lag, *l* = 14. For deaths growth as the outcome, we use *l* = 28. The arguments for choosing these lags are discussed in detail in Appendix E. Alternative lags are explored in Section 4.4.

Estimation equation (1) is based on the structural model of Chernozhukov et al. (2021), who propose the following causal mechanism and identification strategy. First, information *I*_*it*−*l*_ and confounders *W*_*it*−*l*_ are determined at *t* − *l*. Second, policies or NPIs, *P*_*it*−*l*_ are set, given the information and confounders. Third, behaviour *B*_*it*−*l*_ is realized, given the policies, information and confounders in place at that time. It is assumed that behaviour reacts to the information without a significant lag. Finally, the outcome *Y*_*it*_ is realized (with lag *l*) given the policies, behaviour, information and confounders.

By including lagged outcomes, the proposed estimation approach allows for possible endogeneity of the policy interventions *P*_*it*_, that is, the introduction or relaxation of NPIs based on information about the level or the growth rate of cases or deaths. Also, past cases may be correlated with (lagged) government policies or behaviours that are not fully captured by the included policy and behaviour variables.

In Section 4.3 and in Table B19, we also estimate the relationship between policies *P*_*it*_, information, *I*_*it*_ (weekly levels or growth of cases or deaths) and behaviour, *B*_*it*_.

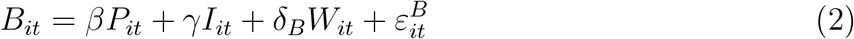

We find strong correlation between policy measures and the Google mobility behavioural proxy in Table B19.

Equation (1) separates the direct effect of policies on outcomes, with the appropriate lag, from the potential indirect effect on outcomes of changes in behaviour captured by the geolocation trends proxy, *B*_*it*−*l*_. In Appendix Table B20, we report estimates without including the behavioural proxy, that is, capturing the total effect of policies on outcomes. Since our estimates of the coefficient *α* in equation (1) are not significantly different from zero, the results without controlling for the behavioural variable *B*_*it*−*l*_ are very similar to those from estimating equation (1).

### 3.2 Data and descriptive analysis

#### Outcomes

Our main outcome of interest is the weekly growth rate of new positive COVID-19 cases, defined below.^12^ We use weekly outcome data to correct for the strong day-of-the-week effect present in COVID-19 data, with markedly lower numbers reported on weekends or holidays. Weekly case growth is a metric that is helpful in assessing trends in the spread of COVID-19, and it has been highlighted in WHO’s weekly epidemiological reports (World Health Organization, 2020).

Specifically, let *C*_*it*_ denote the cumulative case count up to day *t* and define Δ*C*_*it*_ as the weekly COVID-19 cases reported for the 7-day period ending at day *t*:

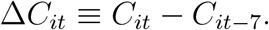

The (log) weekly case growth rate, *Y*_*it*_ is defined as:

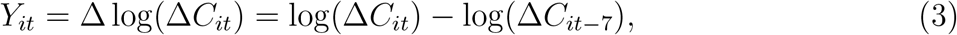

that is, the week-over-week growth in cases in region *i* ending on day *t*.^13^ The weekly test growth rate Δ log(Δ*T*_*it*_) is defined analogously.

#### Policy

In the Ontario analysis, we exploit regional variation in the timing of indoor mask mandates staggered over two months in the province’s 34 PHUs. Fig. 1 displays the gradual introduction of mandates. The exact implementation dates are reported in Table D2. Mandatory indoor masks were introduced first in the Wellington-Dufferin-Guelph PHU on June 12, 2020 and last in the Northwestern PHU on August 17, 2020.^14^

**Figure 1:**
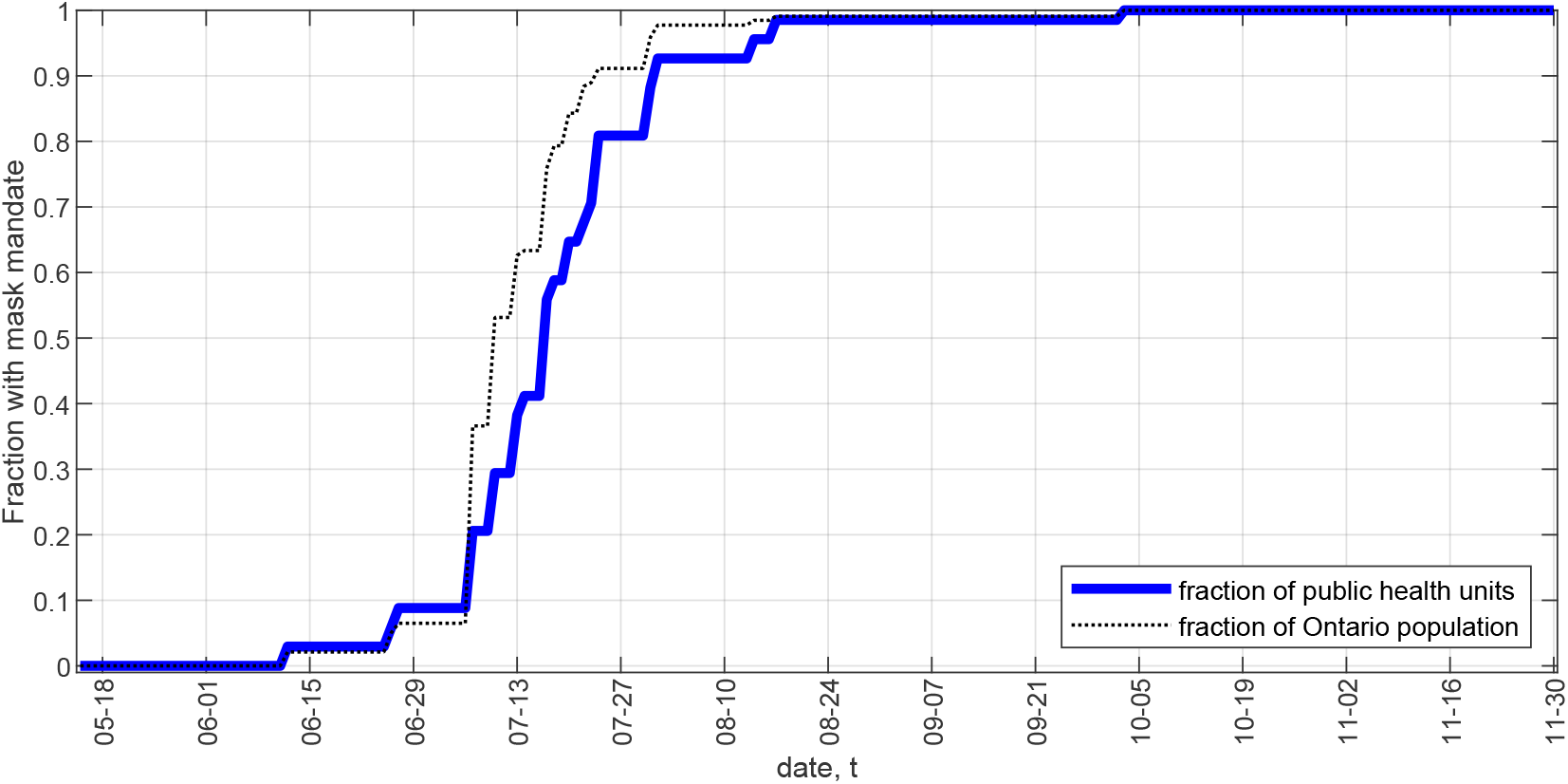
Ontario – mask mandates over time Notes: There are 34 public health units (PHU) in Ontario. See Table D2 for the exact date of mask mandate implementation in each PHU.

We assign numerical values to each of 17 policy indicators, listed and defined in Table D1. The policy variables take values on the interval [0,1], with 0 meaning absence or lowest level of restrictions and 1 meaning most stringent restrictions. A policy value between 0 and 1 indicates partial restrictions, either in intensity (see Table D1 for details and definitions) or in geographical coverage (in large provinces). The policy values are assigned at the daily level for each region, while maintaining comparability across regions. Most non-mask policy indicators are available at the province level only.

Many NPIs were implemented at the same time, both relative to each other and across regions, especially during the March 2020 initial closing-down period.^15^ This causes many of the policy indicators to be highly correlated with each other (see Appendix Table B4). To avoid multi-collinearity problems, we therefore group, via arithmetic averaging, the 17 policy indicators into 5 policy aggregates: (i) *Mask*, which takes value 1 if an indoor mask mandate has been introduced, 0 if not, or value between 0 and 1 if only part of a province has enacted the policy; (ii) *Business/gathering*, which comprises regulations and restrictions on non-essential businesses and retail, personal businesses, restaurants, bars and nightclubs, places of worship, events, gyms and recreation, and limits on gathering; (iii) *School*, which is an indicator of provincial school closure (including Spring and Summer breaks); (iv) *Travel*, which includes international and domestic travel restrictions and self-isolation rules and (v) *Long-term care (LTC)*, which includes NPIs governing the operation of long-term care homes (visitor rules and whether staff are required to work on a single site).

The five policy aggregates are constructed at the daily level and capture both policy restrictions (an increase in the numerical value from 0 toward 1) and policy relaxations (decrease in the numerical value toward zero). In comparison, the policy indicators compiled by Raifman (2020) for the U.S. used in Chernozhukov et al. (2021) are binary “on (1)”/”off (0)” variables.^16^ For consistency with the weekly outcome and information variables and the empirical model timing, we construct the policy aggregates 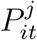 used in the regressions (where *j* = 1, …, 5 denotes policy type) by taking a weekly moving average of the daily policy values, from date *t* − 6 to date *t*.

Fig. 2 plots the values of the five policy aggregates over time for each of the 10 provinces. Travel restrictions, school closures and business closures were initially implemented in a relatively short period in the middle of March 2020. There is variation in the *Travel* aggregate since some Canadian provinces (the Atlantic provinces) implemented strict inter-provincial domestic travel and self-isolation restrictions in addition to the federal regulations regarding international travel. Restrictions on long-term care facilities were introduced more gradually, with large variation across the provinces. In the period May-November 2020, there is more policy intensity variation, especially in the business and gatherings category, as the different provinces implemented their own ‘re-opening’ plans and strategies. Mask mandates were introduced in Ontario starting from June in a few smaller PHUs and in early July in the most populous PHUs, Toronto, Ottawa and Peel (see Table D2). In Quebec, indoor masks were mandated province-wide on July 18 while other provinces such as British Columbia and Prince Edward Island did not introduce mask mandates until November (see Table D3 for the complete list of dates).

**Figure 2:**
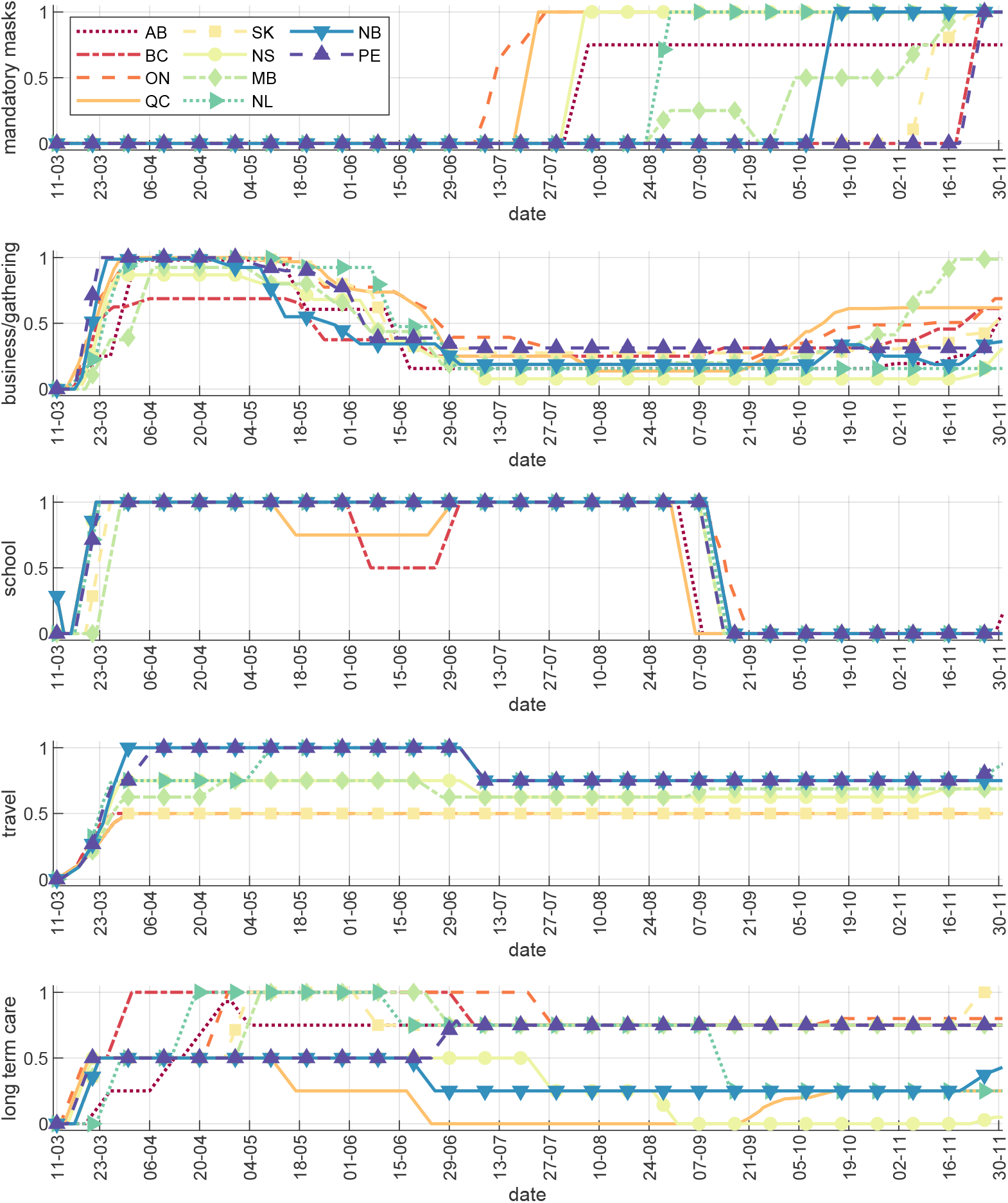
Policy aggregates – Canada Notes: The figure plots the numerical values of the five policy aggregates (Mask, Business/gathering, School, Travel and Long-term care, LTC) over time, for each of the 10 provinces. The mask policy values for Ontario (ON) reflect the gradual adoption of mandates (see Fig. 1) and the respective PHUs’ population sizes.

There are two empirical challenges specific to the Canadian context and data. The first is the presence of small provinces or sub-regions with very few COVID-19 cases or deaths. In Section 4.4, we perform robustness checks using different ways of handling the observations with very few cases (in particular zero cases). The second data limitation is that there are only 10 provinces in Canada and 34 public health units in Ontario, compared to 51 U.S. states in Chernozhukov et al. (2021). To account for the resulting small number of clusters in the estimation, we compute and report wild bootstrap standard errors and p-values, as proposed by Cameron et al. (2008).^17^ On the other hand, our data has the advantage of a longer time horizon (March to November) and non-binary, more detailed policy variables compared to Raifman et al. (2020).

#### Behaviour proxy

We follow Chernozhukov et al. (2021) and other authors in interpreting the location change indices from the Google Community Mobility reports as proxies for changes in people’s behaviour during the pandemic, keeping in mind that location is only one aspect of behaviour relevant to the spread of COVID-19. The general pattern in the data (see Fig. C3) shows sharply reduced frequency of recorded geo-locations in shops, workplaces and transit early in the pandemic (March), with a subsequent gradual increase back toward the baseline (except for transit), a flattening out in July and August, and a relatively small decline in the Fall months.

Several of the six available location indicators (retail, grocery and pharmacy, workplaces, transit, parks and residential) are highly correlated with each other (see Tables B1 and B2) and/or contain many missing observations for the smaller PHUs and provinces. To address these data limitations and the possible impact of collinearity on the estimation results, we use as proxy for behavioural changes the arithmetic average of three mobility indicators: “retail”, “grocery and pharmacy” and “workplaces”.^18^ To be consistent with the weekly outcome variables and to mitigate day-of-the-week geo-location variation, we construct the Behaviour proxy *B*_*it*_ by taking a weekly moving average of the 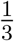 (retail + grocery and pharmacy + workplaces) data, from date *t* − 6 to date *t*.^19^ As a result, our empirical analysis uses weekly totals (for cases, tests and deaths) or weekly moving averages (for policies and the behaviour proxy) for all variables recorded on daily basis.

Tables B3 and B4 display the correlation between the behaviour proxy *B*_*it*_ and the five policy aggregates 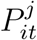. The behaviour proxy and mask mandate variable are not highly correlated, suggesting that the effect of mask mandates on COVID-19 outcomes is likely not dependent on location behaviour changes.

#### Information

We use the weekly cases and case growth variables defined above, Δ*C*_*it*_ and *Y*_*it*_, to construct the information variables *I* used in estimation equation (1). Specifically, we use as information the lagged weekly case growth rate, Δ log(Δ*C*_*it*−*l*_) and the log of past weekly cases, log(Δ*C*_*it*−*l*_). We also use the lagged provincial (in the Ontario analysis) or national (in the Canada analysis) case growth rate and the log of weekly cases as additional information variables in some specifications. A two-week information lag, *l* = 14, is used in the baseline analysis. In the supplementary analysis using the death growth rate as the outcome, we use information on past deaths and a four-week lag (see Appendix A).

#### Control variables

In all regressions, we control for region fixed effects (PHU or province) and the weekly COVID-19 tests growth rate Δ log(Δ*T*_*it*_), where *T*_*it*_ denotes cumulative tests in region *i* until date *t* and Δ*T*_*it*_ is defined analogously to Δ*C*_*it*_.^20^ Our baseline specification includes week fixed effects, and we also report results with no time trend and with cubic trend in days in Appendix B. In robustness checks, we also include news or weather variables as controls (see Section 4.4).

#### Time period

We use the period May 15 to November 30, 2020 for our analysis with Ontario PHU data and the period March 11 to November 30, 2020 for the analysis with provincial data. The start date for the Ontario sample (May 15) is chosen as approximately two weeks after the last Spring restrictions were implemented and four weeks before the first PHU mask mandate was introduced. Sensitivity checks with different initial dates (May 1, June 1 and June 15) are reported in Section 4.4, with our main results remaining robust. The initial date for the Canada sample (March 11) was chosen as the first date on which each province reported at least one COVID-19 test (so that cases could be registered). Alternative initial dates are also considered in Section 4.4. We chose November 30 as the end date to avoid possible confounding effects from the holidays season and, given that the main focus of the paper is mask mandates, because by November mask usage was already very high in both jurisdictions with and without mandates (see Fig. C2).

## 4 Results

### 4.1 Mask mandates in Ontario public health regions

Fig. 3 displays results from an event study analysis of PHU mask mandates and the change in case growth rate in Ontario from six weeks before to five weeks after the mask mandate, where *T* = 0 is the mandate implementation date and the reference point is one week before the implementation of the mask mandate (*T* = –1).

**Figure 3:**
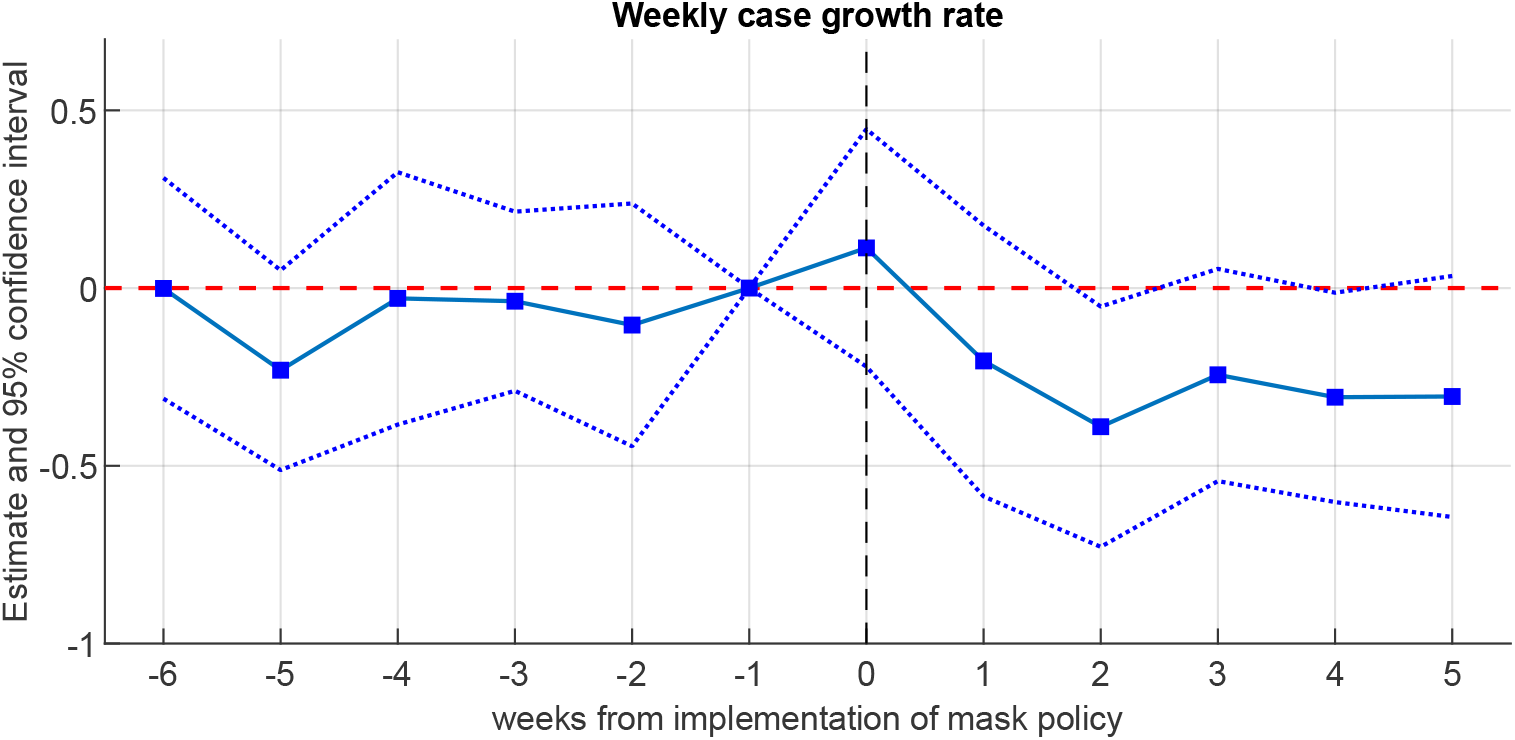
Ontario – Event study of weekly case growth Notes: Time period – May 15 to November 30, 2020. The outcome variable is the weekly case growth rate *Y*_*it*_ = Δ log(Δ*C*_*it*_). The figure plots the estimates from a version of equation (1) where the mask policy variable is replaced by the interaction of being in the ‘treatment’ group (imposed mask mandate) with a series of dummies for each week, ranging from exactly 6 weeks before (*T* = −6) to exactly 5 weeks after the mandate (*T* = +5), where *T* = 0 is the mandate implementation date. The reference point is 1 week before the implementation (*T* = –1). Wild bootstrap (cgmwildboot) standard errors clustered by province with 5,000 repetitions are used to construct the confidence intervals.

We first note that Fig. 3 does not exhibit a pre-trend – the estimates are close to and statistically indistinguishable from zero before the PHU mask mandates are implemented. This helps address the potential concern that PHUs that implemented mask mandates at different times may have had a different trend in case growth. Second, we see a slight uptick at *T* = 0 (day 0 to 6), which may suggest that (some) mask mandates were implemented when case growth was high, a potential endogeneity concern. However, the *T* = 0 estimate is economically small and not statistically different from zero. Third, we confirm that the effect of mask mandates on case growth entails a time lag: we first observe a decrease in case growth at *T* = +1 (day 7 to 13), while a larger and statistically significant decrease occurs at *T* = +2 (day 14 to 20) justifying the 14-day lag used in our main specification (1). Fourth, the mask mandate effect appears persistent rather than transitory, since the reduction in case growth after *T* = +2 does not revert to its level from before *T* = 0.

Table 1 shows the estimates of equation (1), in which we control for other policies, behaviour and information, as explained in Section 3.1.^21^ We report wild bootstrap p-values clustered at the PHU level to account for the small number of clusters.^22^ Column (1) in Table 1 uses lagged cases and lagged case growth at the PHU level as information; column (2) also includes lagged cases and lagged case growth at the province level as additional information variables.^23^ We also include week fixed effects as a flexible way to control for additional province-wide factors possibly affecting the spread of COVID-19, e.g., income support policies, adaptation to the pandemic over time, so-called ‘COVID fatigue’, etc. All robustness checks use this specification. In Appendix Table B5, we also estimate (1) without a time trend or using a cubic time trend in days from the beginning of the sample.

**Table 1:**
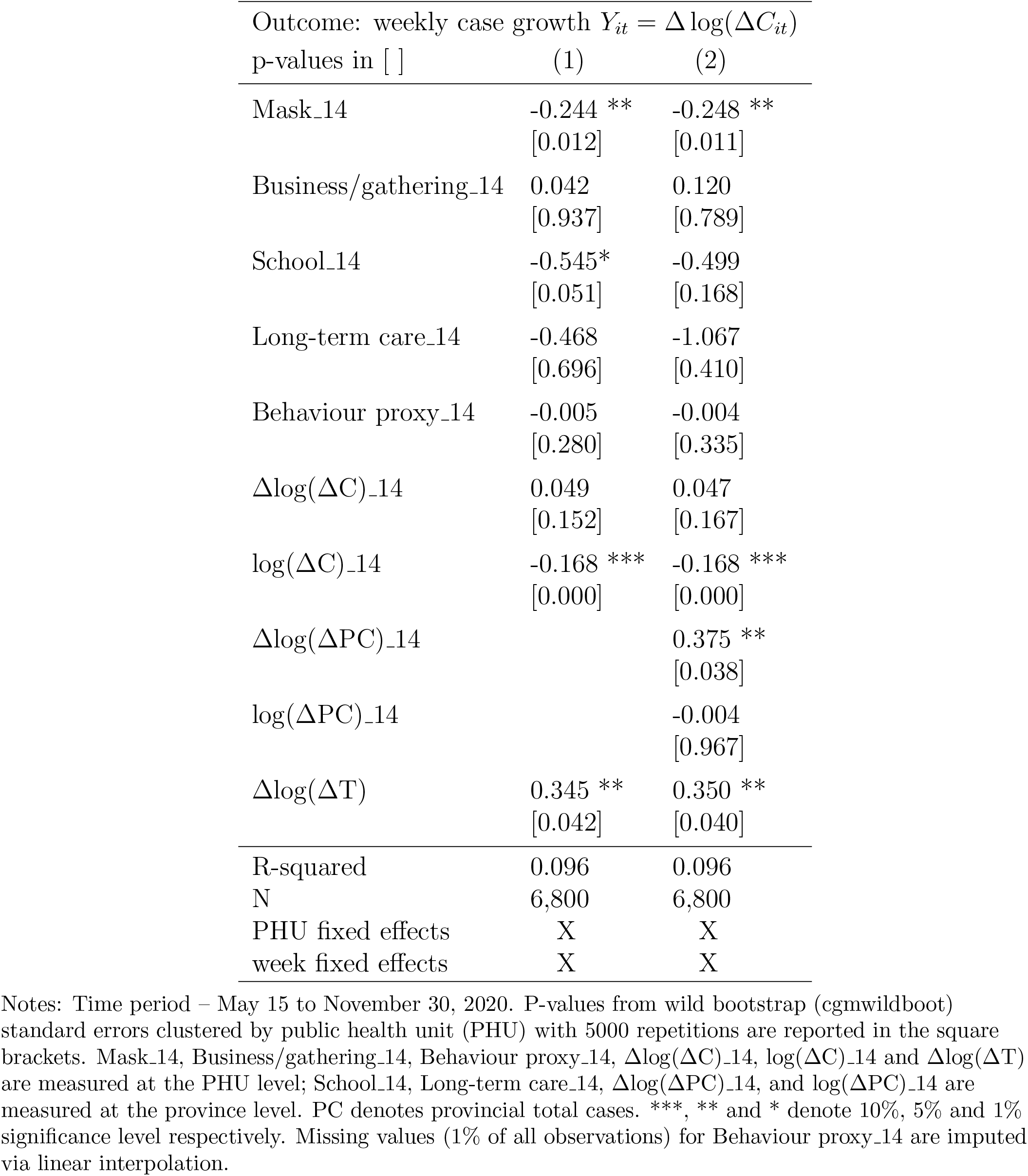
Ontario – Main Results

The estimates in Table 1 imply that, controlling for other policies, information, testing, and geo-location behaviour, mandatory indoor face masks are associated with a decrease of 24–25 log points (*p* < 0.02) in the weekly case growth rate, two weeks after implementation. This result can be interpreted as a 22% weekly reduction in new cases, relative to the trend without mask mandate.^24^ The magnitude of the mask policy estimates is not very sensitive to whether lagged province-level data are included as additional information. We do not find a consistent statistically significant effect of the other NPIs in the Ontario sample.

The results in Table 1 indicate that indoor mask mandates have been a powerful preventive measure in the COVID-19 context. Our estimates of the mandates’ impact in Ontario’s PHUs are larger than the 10 percentage point reduction in case growth estimated by Chernozhukov et al. (2021) for the U.S. A possible explanation is that Ontario’s mask mandates are more comprehensive: we evaluate the effect of universal indoor mask-wearing for the public at large rather than the effect of mask-wearing for *employees only* in Chernozhukov et al. (2021). Differences in the compliance rate may also be a factor; we discuss this potential mechanism in Section 4.3.

Table 1 also shows a statistically significant negative association between information (log of past cases, log(ΔC) 14), and current weekly case growth (*p* < 0.001 in all specifications). This indicates that a higher (lower) level of cases two weeks prior is associated with lower (higher) current case growth, which is consistent with a possible behavioural feedback. Therefore, while the location-based proxy *B*_*it*_ allows for certain behavioural responses, it may not capture other important aspects of behaviour (e.g., frequent hand-washing, physical distancing or the rate of social interactions) that can be influenced by the observed case counts. In fact, our coefficient estimate on the behaviour proxy *B*_*it*_ is not statistically significantly different from zero (both in Table 1 and in the province-level Table 2 below), unlike in Chernozhukov et al. (2021). In Table B19, we find strong contemporaneous correlations between policy measures, log cases, and the Google mobility behavioural proxy from estimating equation (2). This suggests that the information (lagged cases) and the lagged policy variables in (1) may absorb lagged behavioural responses proxied by *B*_*it*−*l*_ or other latent behavioural changes not captured by *B*_*it*−*l*_. Finally, as expected, the weekly growth rate of COVID cases is positively correlated with the weekly tests growth rate.

**Table 2:** Canada – Main Results

| Outcome: weekly case growth $Y_{it} = \Delta \log(\Delta C_{it})$ | | |
| --- | --- | --- |
| p-values in [ ] | (1) | (2) |
| Mask_14 | -0.219 **<br>[0.028] | -0.223 **<br>[0.025] |
| Business/gathering_14 | -0.580 *<br>[0.068] | -0.594 *<br>[0.076] |
| School_14 | -0.148<br>[0.242] | -0.162<br>[0.182] |
| Travel_14 | 0.206<br>[0.705] | 0.161<br>[0.764] |
| Long-term care_14 | -0.093<br>[0.672] | -0.097<br>[0.665] |
| Behaviour proxy_14 | -0.005<br>[0.763] | -0.004<br>[0.788] |
| $\Delta \log(\Delta C)_14$ | -0.047<br>[0.527] | -0.050<br>[0.519] |
| $\log(\Delta C)_14$ | -0.170 **<br>[0.043] | -0.172 **<br>[0.040] |
| $\Delta \log(\Delta NC)_14$ | | 0.224<br>[0.327] |
| $\log(\Delta NC)_14$ | | 0.184 **<br>[0.046] |
| $\Delta \log(\Delta T)$ | 0.295 ***<br>[0.000] | 0.297 ***<br>[0.000] |
| R-squared | 0.338 | 0.339 |
| N | 2,650 | 2,650 |
| province fixed effects | X | X |
| week fixed effects | X | X |

Notes: Time period – March 11 to November 30, 2020. P-values from wild bootstrap (cgmwildboot) standard errors clustered by province with 5,000 repetitions are reported in the square brackets. NC denotes national total cases. \*\*\*, \*\* and \* denote 10%, 5% and 1% significance level respectively.

Fig. 4 displays the estimates on the mask mandate variable Mask 14 from equation (1) for different sample end dates, compared to the baseline results for November 30 in Table 1. The results show that the estimated mandate effect is stable over these dates, as all PHU-level mandates were implemented by August 17.

**Figure 4:**
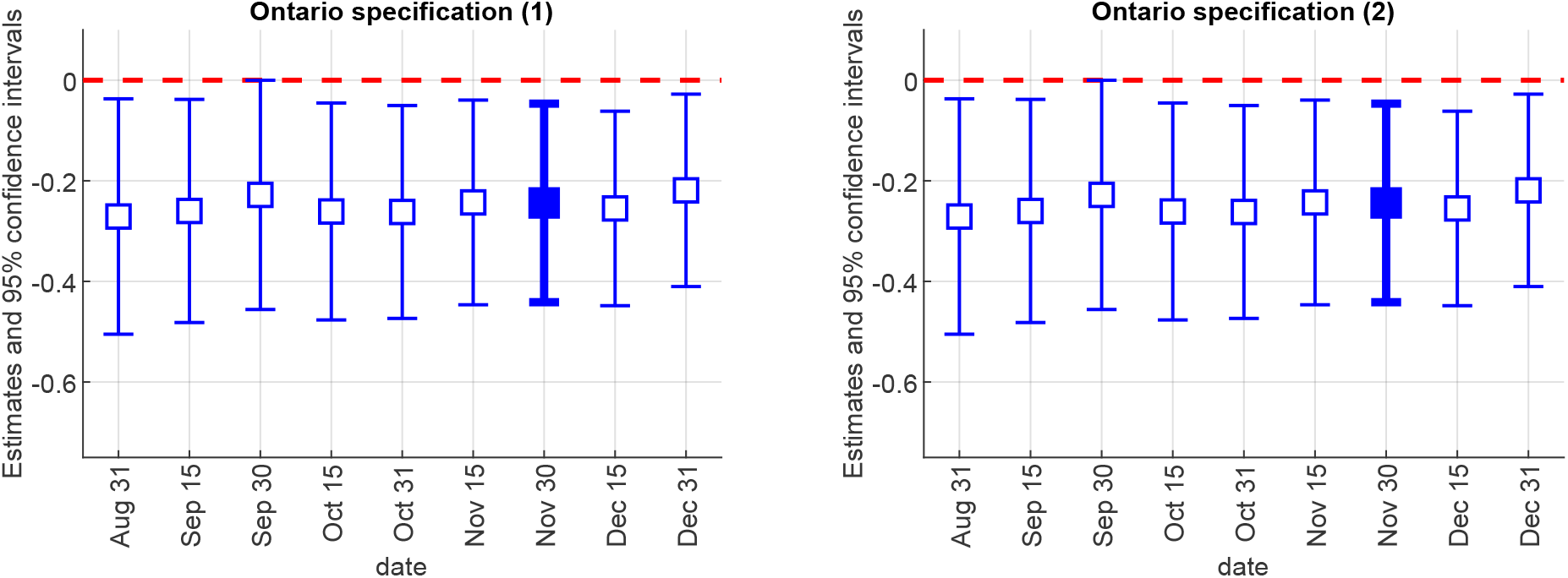
Ontario – Mask mandate estimate by date Notes: The figure plots the coefficient estimates on Mask 14, with 95% confidence intervals, from equation (1) for different end dates of the sample. The baseline specifications (Table 1) use November 30. The left panel corresponds to column (1) in Table 1; the right panel corresponds to column (2) in Table 1.

### 4.2 Province-level results

We next evaluate the impact of NPIs on COVID-19 case growth in Canada as a whole, by using variation in the timing of policy interventions across the 10 provinces. We also examine NPIs for which there is no data variation across Ontario’s PHUs (e.g., travel or long-term care regulations), in addition to mask mandates.

Table 2 displays the estimates of equation (1) for weekly case growth, along with wild bootstrap p-values, clustered at the province level (see Table B10 for other methods of computing the standard errors). Column (1) uses lagged cases and lagged case growth at the province level as information while column (2) additionally includes lagged cases and case growth at the national level. Both specifications include province and week fixed effects (see Table B5 for different time controls).

The most robust result in Table 2 is the estimate on mask mandates (Mask 14). We find that mask mandates are associated with a significant reduction in weekly case growth of 22 log points (p-values < 0.03), which corresponds to a 20% weekly reduction in new cases, relative to the trend in absence of mandate. It is reassuring that these province-level results are very close in magnitude and consistent with our results using Ontario PHU data.

Table 2 further suggests that restrictions on businesses and gatherings are associated with a reduction in weekly case growth of 58–59 log points or, vice versa, that relaxing business/gathering restrictions is associated with higher case growth. These estimates correspond to a 44–45% weekly decrease in new cases, relative to the trend in absence of mandate. We obtain similar results in robustness Tables B5, B12 and B16. Note, however, that the business/gathering estimates are noisier than our estimates for mask mandates (p-values < 0.08) and lose statistical significance in some robustness checks (Table B9). Still, these results suggest that relaxed restrictions and the associated increase in business and work-place activity or gatherings (including restaurants, bars and retail) may be an important offsetting factor for the estimated impact of mask mandates on COVID-19 case growth.

As in Table 1, we again find that weekly case growth is negatively associated with weekly cases two weeks prior (log(ΔC) 14 in Table 2) and positively associated with the weekly tests growth, Δ log(Δ*T*) (p-value < 0.001).^25^

Fig. 5 displays the estimates of the coefficient on the mask mandate variable Mask 14 in equation (1) for different sample end dates, compared to the baseline results for November 30 in Table 2. The figure shows that the estimated mask mandate effect slowly diminishes over time, from −0.35 until Sep. 15 to −0.17 until Dec. 31. A possible explanation is the steady increase in self-reported mask usage in the provinces without mandates over this period (see Fig. C2), which is also consistent with the declining estimate of the effect of mask mandates on mask usage over time shown in Fig. 7 in Section 4.3.

**Figure 5:**
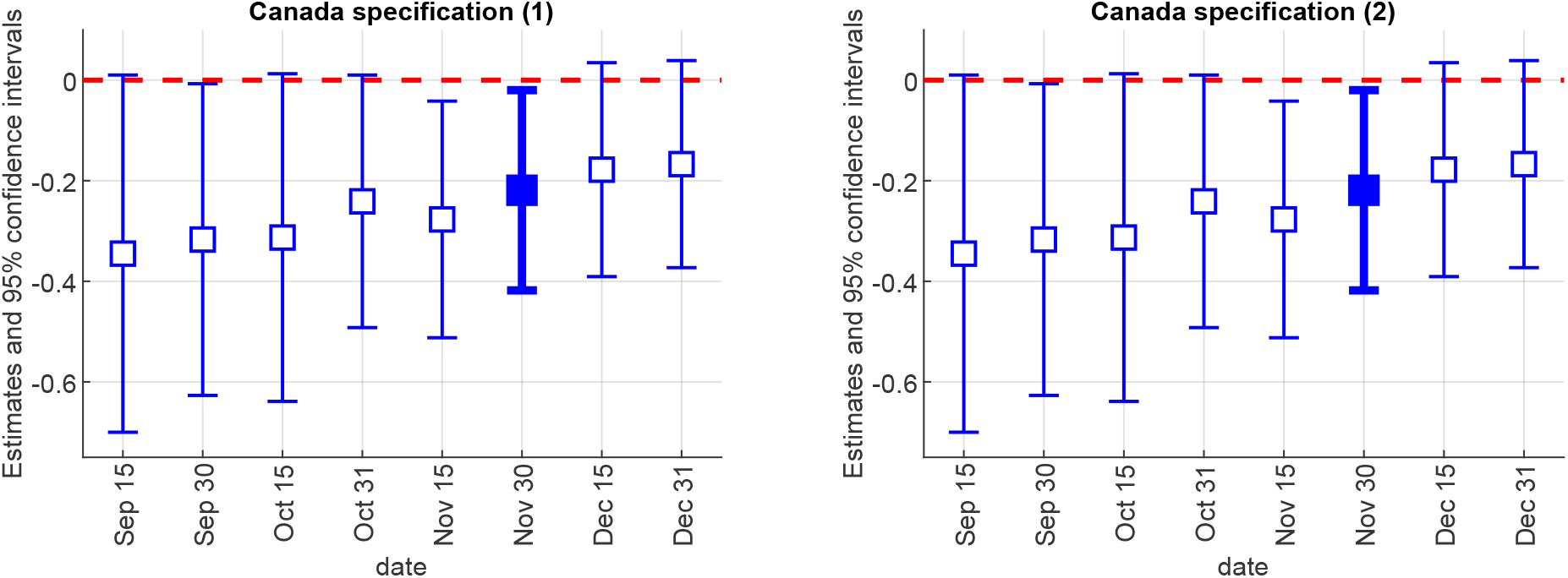
Canada – Mask mandate estimate by date Notes: The figure plots the coefficient estimates on Mask 14, with 95% confidence intervals, from equation (1) for different end dates of the sample. The baseline specifications (Table 2) use November 30. The left panel corresponds to column (1) in Table 2; the right panel corresponds to column (2) in Table 2.

**Figure 6:**
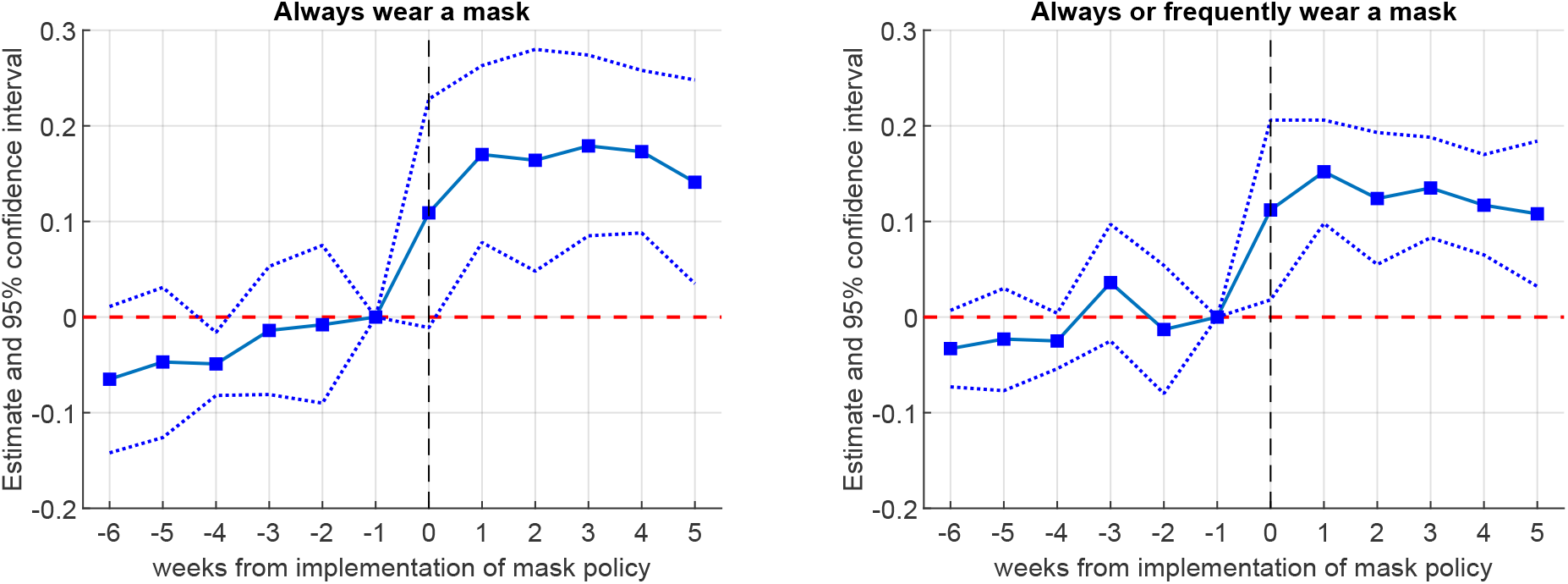
Event study of self-reported mask usage in Canada Notes: Time period – April 2 to November 30, 2020. The data source is YouGov. The binary outcome variable takes value 1 if the respondent answered “Always” (in the left panel) or “Always” or “Frequently” (in the right panel) to “Thinking about the last seven days, how often have you worn a face mask outside your home?” The figure plots the estimates from a version of equation (2) where the mask policy variable is replaced by the interaction of a variable denoting being in the ‘treatment group’ (imposed mask mandate) with a series of dummies for each week, ranging from exactly 6 weeks before the mask mandate (*T* = −6) to exactly 5 weeks after the mandate (*T* = +5), where *T* = 0 is the mandate implementation date. The reference point is 1 week before the implementation (*T* = –1). Wild bootstrap (cgmwildboot) standard errors clustered by province with 5,000 repetitions are used to construct the confidence intervals. Sample weights are used.

**Figure 7:**
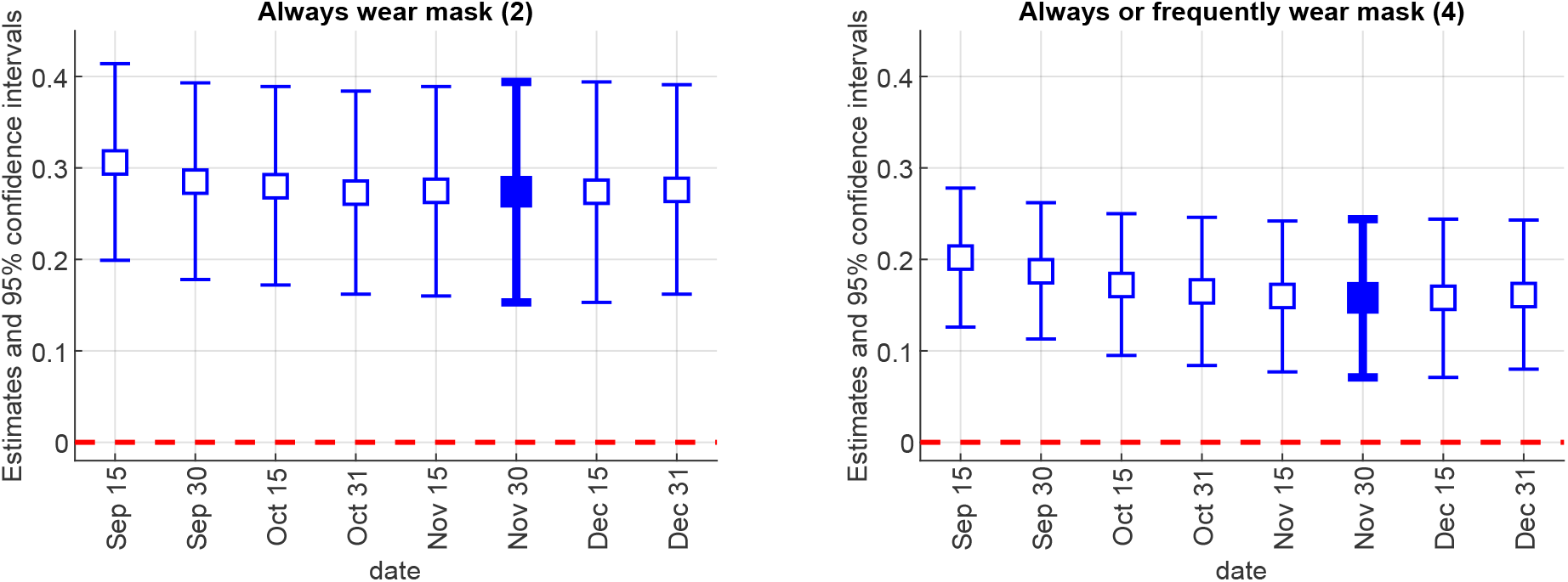
Mask usage estimate by date Notes: The figure plots the coefficient estimates on Mask, with 95% confidence intervals, from equation (2) for different end dates of the sample. The baseline specifications (Table 3) use November 30. The left panel corresponds to column (2) in Table 3; the right panel corresponds to column (4) in Table 3.

### 4.3 Mask usage

The effectiveness of any public policy critically depends on whether and how it affects people’s behaviour. In this section, we use self-reported survey data on mask usage to examine whether mask mandates increased mask use in Canada (“first-stage” analysis).

We use data from the YouGov COVID-19 Public Monitor (Jones et al. 2021), which consists of multiple waves of public opinion surveys fielded regularly since early April 2020 in many countries.^26^ Here, we focus on inter-provincial comparisons within Canada. Our main variable of interest is the response to the question “Thinking about the last 7 days, how often have you worn a face mask outside your home (e.g. when on public transport, going to a supermarket, going to a main road)?” The possible answers are “Always”, “Frequently”, “Sometimes”, “Rarely”, and “Not at all”. We create a binary variable taking value 1 if the response is “Always” and 0 otherwise, and another variable taking value of 1 if the respondent answered either “Always” or “Frequently” and 0 otherwise.

We begin with an illustration of self-reported mask usage in Canada from April to November 2020. Fig. C2 plots the average self-reported mask usage (response “Always”) in the provinces with and without mask mandates.^27^ The figure shows that self-reported mask usage is higher, by up to 50 percentage points in early August, in the provinces with a mask mandate compared to the provinces without a mandate. Since Fig. C2 does not account for compositional changes in the data and other factors, we formally estimate equation (2), using self-reported mask usage as the behavioural outcome.^28^

Fig. 6 displays the results from an event study analysis of mask mandates and the change in mask usage, ranging from 6 weeks before the mask mandate to 5 weeks after the mask mandate (*T* = –6 to +5, where *T* = 0 is the implementation date of the mask mandate). The reference point is one week before the implementation of the mask mandate (*T* = –1). The left and right panels of Fig. 6 present results for the “Always” and “Always” or “Frequently” mask usage responses, respectively.

We make several observations. First, neither panel shows a pre-trend – the estimates are statistically indistinguishable from zero before the mask mandates are implemented. This addresses the potential concern that provinces that implemented mask mandates may have had a different trend in mask usage than provinces that did not. Second, the effect of mask mandates on mask usage is immediate: an increase of roughly 11 percentage points on average, as soon as the mask policy is implemented (*T* = 0). Third, the effect appears persistent rather than transitory, since mask usage does not revert to its level before *T* = 0.

Table 3 displays the estimates on self-reported mask usage in equation (2) along with wild bootstrap p-values clustered at the province level. As in Tables 1 and 2, our baseline specification uses week fixed effects.^29^ The results show that mask mandates are associated with 27 percentage point increase in self-reported mask usage, for the response “Always” (*p* < 0.001), from a base of self-reported mask usage without mask mandate of 33.2%. Similarly, “Always” or “Frequent” mask usage increases by 16 percentage points.^30,31^

**Table 3:** Canada – Self-reported mask usage

| p-values in [ ] | Outcome: Wear mask |  |  |  |
| --- | --- | --- | --- | --- |
|  | “Always”<br>(1) | “Always”<br>(2) | “Always” or “Frequently”<br>(3) | “Always” or “Frequently”<br>(4) |
| Mask | 0.273 ***<br>[0.000] | 0.274 ***<br>[0.000] | 0.158 ***<br>[0.000] | 0.158 ***<br>[0.000] |
| $\Delta\log(\Delta C)$ | -0.005<br>[0.680] | -0.007<br>[0.599] | -0.016<br>[0.183] | -0.018<br>[0.170] |
| $\log(\Delta C)$ | 0.004<br>[0.585] | 0.005<br>[0.561] | 0.015 **<br>[0.010] | 0.015 **<br>[0.019] |
| $\Delta\log(\Delta NC)$ | | 0.129<br>[0.356] | | 0.137<br>[0.372] |
| $\log(\Delta NC)$ | | -0.073<br>[0.298] | | 0.033<br>[0.663] |
| R-squared | 0.274 | 0.274 | 0.254 | 0.255 |
| N | 15,808 | 15,808 | 15,808 | 15,808 |
| mean w/o mask mandate | 0.332 | 0.332 | 0.507 | 0.507 |
| individual characteristics | X | X | X | X |
| province fixed effects | X | X | X | X |
| week fixed effects | X | X | X | X |

These results show that mask mandates exhibit broad compliance in Canada and establish a possible mechanism for the significant impact of the mandates on the spread of COVID-19 that we find. That said, given that mask mandates do not change everyone’s behaviour, our estimates in Tables 1 and 2 represent intent-to-treat effects. The full effect of the entire population shifting from not wearing to wearing masks is likely larger.^32^

There has been a heated debate on whether mask wearing may create a false sense of security and reduce adherence to other preventive measures. We investigate this question using the YouGov survey data. As Tables B14 and B15 in the Appendix indicate, we find no evidence that mask mandates in Canada had an offsetting effect on other preventive measures such as hand washing, using sanitizer, avoiding gatherings, and avoiding touching objects in public during the period we study.^33^

Finally, in Fig. 7, we plot the estimates for the mask mandate variable from Table 3 for different dates. The estimates slightly decrease over time, which is consistent with the slowly diminishing effect of the mask mandates on case growth in Canada shown in Fig. 5.

### 4.4 Robustness

#### Zero weekly cases

A possible concern may be that the dependent variable *Y*_*it*_ = Δ log(Δ*C*_*it*_) is not well defined when the weekly case totals Δ*C*_*it*_ or Δ*C*_*it*−7_ equal zero. As in Chernozhukov et al. (2021), we replace log(0) with −1 in our baseline specification.^34^ We check the robustness of our estimates to alternative treatments of zero weekly cases in Appendix Tables B6 and B9 – replacing log(0) with 0, adding 1 to Δ*C*_*it*_ before taking logs, or population-weighted least squares. Our main results on mask mandates in Ontario PHUs and Canadian provinces are robust to these alternative specifications, with the single exception of columns (7) and (8) in Table B6 where the mask mandate estimate loses statistical significance at the 10% level due to reduced power.

#### Policy collinearity

A possible concern is that some NPIs (e.g., international travel restrictions or school closures) were implemented within a very short time interval. Thus, we may lack sufficient regional variation to distinguish and identify the separate effect of each policy.^35^ Collinearity in the policy aggregates could also affect the standard errors and the signs of the estimated coefficients. Table B8 for Ontario and Table B11 for Canada in the Appendix report the estimates from our baseline specification when omitting one policy at a time or when including the mask mandate policy only. Our mask mandate estimates remain largely robust, with the exception of the last columns in Table B11 where the estimates’ statistical significance is affected. This confirms the importance of controlling for all NPIs.

#### Alternative initial dates

Fig. C4 and C5 show that our estimates and confidence intervals for the effect of mask mandates on case growth in Ontario and Canada do not vary much with the initial date of the respective samples.

#### Alternative lags

We also check alternative time lags, of either shorter or longer duration, centered around the baseline value of 14 days. Fig. C6 for Ontario and Fig. C7 for Canada plot the estimates and 95% confidence intervals. The mask mandate estimates for Canada remain fairly consistent for the different plausible lag values while the Ontario sample estimates are a bit more sensitive to the lag length, with our baseline estimates being in the middle in terms of magnitude.

#### Omitted variables

The Google mobility behaviour proxy variable likely misses some aspects of people’s behaviour that could be relevant for COVID-19 transmission. One such factor could be weather. For example, good weather may cause more people to spend time outside and lower the chance of viral transmission. Columns (3) and (4) in Table B12 report estimates for Canada including lagged weather variables (daily maximum and minimum temperatures and precipitation for the largest city in each province^36^) as additional controls. The estimates on mask mandates remain statistically significantly negative (p-value < 0.03) but are slightly reduced compared to the baseline values in columns (1)–(2). While we find no effect of weather on case growth, we note that any possible effect of seasonal change would be absorbed by the week fixed effects, and any possible effect of day-to-day weather may be obscured by the stochastic lag between transmission and detection.

Another possible concern is that the information variables (lagged cases and lagged case growth) may not fully capture the actual information based on which people react or adjust their behaviour, possibly affecting the observed case growth. Columns (5) and (6) in Table B12 add a national-level “News” variable to our baseline specification, defined as the number of daily search results for “coronavirus” or “COVID-19” from the *ProQuest Canadian Newsstream* news aggregator service (see Appendix D for more details). In column (6), the news variable is statistically significant at the 10% level. Our estimates on mask mandates and business/gathering remain very close to those in the baseline.

#### Heterogeneous treatment effects

We address the possibility of heterogeneous treatment effects in two ways. First, several recent papers point out that the two-way fixed effects estimator (week and PHU/province fixed effects in our case) is a weighted average of treatment effects, where some of the weights can be negative if the treatment effect is not constant across groups and over time.^37^ In particular, de Chaisemartin and D’Haultfoeuille (2018) develop an estimator that is valid under these conditions. We apply their estimator to our main analysis, regarding case growth in Ontario.^38^ We obtain a mask mandate estimate of −0.236 (s.e. 0.415), which is very close in magnitude to our two-way fixed effect estimates (−0.244 and −0.248) in Table 1.^39^

Second, we also compute and compare mask the mandate estimates for ‘early’ vs. ‘late’ PHU adopters in Ontario. Specifically, we define as ‘early’ adopters the PHUs that implemented mask mandates before July 15 (nearly half of the PHUs adopted mask mandates by then), and ‘late’ adopters as the PHUs that implemented mandates after July 15. We obtain similar estimates for the early and late adopters (−0.266 and −0.255).^40^ Jointly, these results suggest that the effects of mask mandates on case growth are unlikely to be heterogeneous across regions and time in a way that affects our estimates, which is also consistent with the estimates’ stability over different dates on Fig. 4.

### 4.5 Counterfactuals

We use our estimates from Tables 1 and 2 to evaluate several counterfactuals in which the actual mask policy is replaced by an alternative hypothetical policy. Letting *t*_0_ be the implementation date of a counterfactual policy, we set the counterfactual weekly case count, 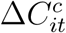, equal to Δ*C*_*it*_ for all *t* < *t*_0_. For each date *t* ≥ *t*_0_, using the definition of *Y*_*it*_ in (3), we compute 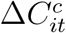 and the counterfactual weekly case growth rate, 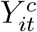, as follows:

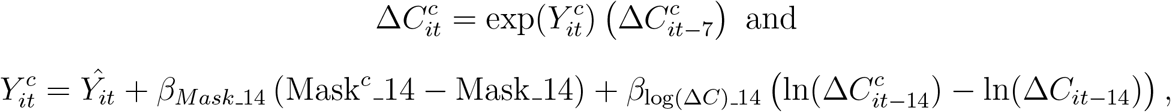

where *Ŷ*_*it*_ is the regression-fitted value of weekly case growth; *β*_*Mask* 14_ is the coefficient estimate (−0.248 or −0.223) on the mask mandate variable Mask 14 in baseline specification, column (2) in Table 1 or 2, depending on the counterfactual; Mask^*c*^ 14 is the counterfactual policy (e.g., different implementation date, wider geographic coverage or absence of mask mandate); and *β*_log(Δ*C*)_14_ is the coefficient estimate (−0.168 or −0.172) on lagged cases log(Δ*C*) 14 in Table 1 or 2, column (2). The coefficient *β*_log(Δ*C*)_14_ adjusts the counterfactual case growth rate for the negative statistically significant association between the weekly case total two weeks prior and time-*t* case growth. This feedback effect may be due to people being more careful when they perceive the risk of infection to be higher.

Fig. 8 and 9 show results from two counterfactual policy evaluations. The first counterfactual, depicted in the left panels, assumes that masks are adopted everywhere at the earliest date observed in the data (June 12, 2020). Fig. 8 shows that if mask mandates were implemented in all Ontario PHUs at the earliest observed date, this could have led to an average reduction of 770 cases per week over the simulation period Jun. 26 to Nov. 30, holding all else equal. The estimated reduction is in the range 70 to 500 cases per week early on, in July–August, peaks at about 2,200 cases per week in mid-October, and decreases to under 1,000 cases per week by the end of November. For Canada as a whole, a country-wide adoption of mask mandates in June is estimated to reduce new weekly COVID-19 cases by 22% on average over the simulation period, which amounts to a total reduction of over 50,000 cases. The estimated average weekly reduction is by about 680 cases for July–August and peaks at 9,600 cases per week in November. In both simulations, the feedback effect via *β*_log(Δ*C*)_14_ (lagged cases as information) starts moderating the decrease in cases two weeks after the initial impact of the counterfactual policy.

**Figure 8:**
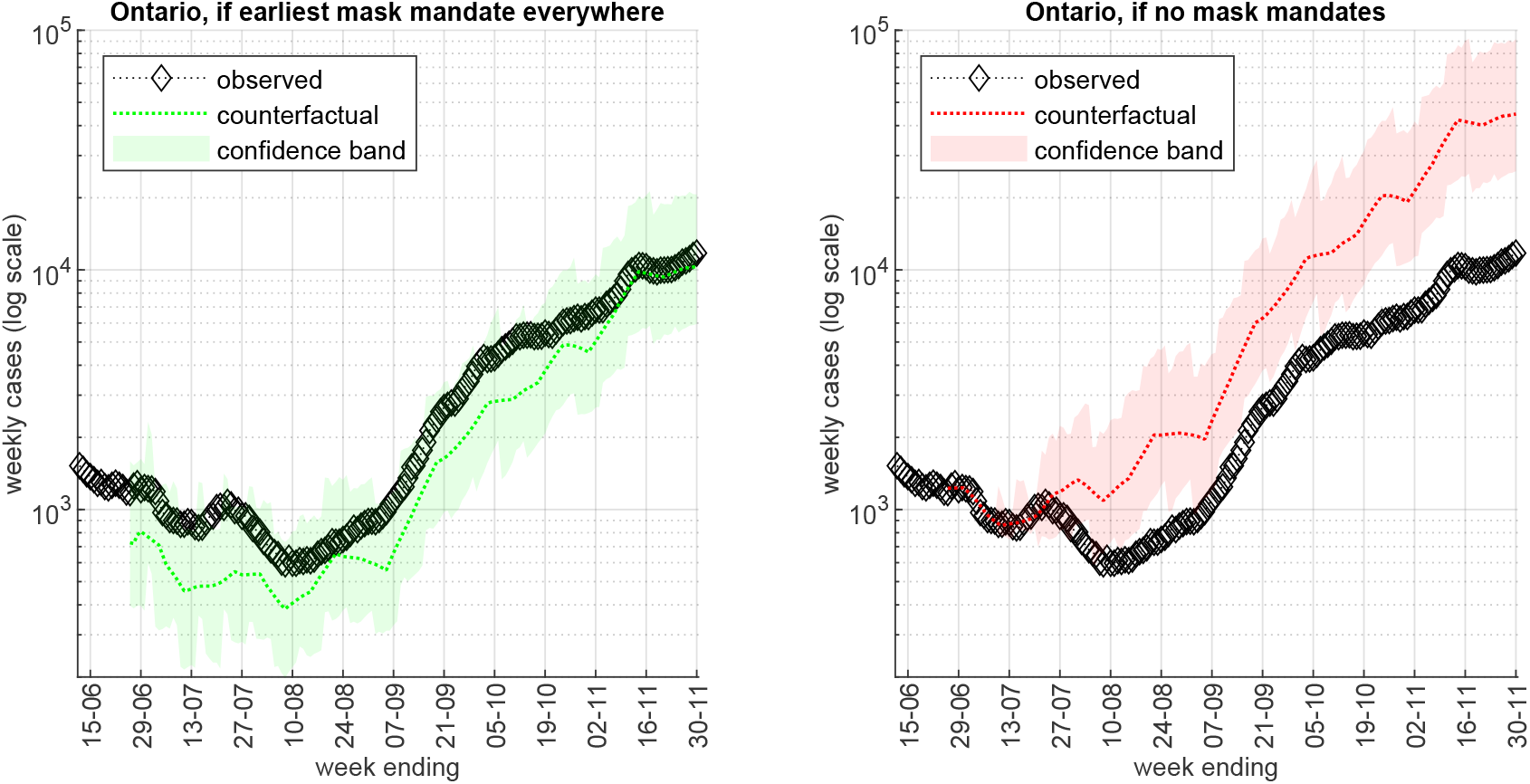
Ontario – Counterfactuals Notes: The left panel assumes that mask mandates were adopted in all PHUs on June 12, 2020 (date of the earliest mask mandate in Ontario). The right panel assumes that mask mandates were not adopted in any PHU. We use the estimates from column (2) of Table 1. The diamonds plot observed weekly cases from *t* −6 to *t*, the dotted lines plot the 7-day moving average of the counterfactual mean value, and the shaded areas are 5-95 percentile confidence bands.

**Figure 9:**
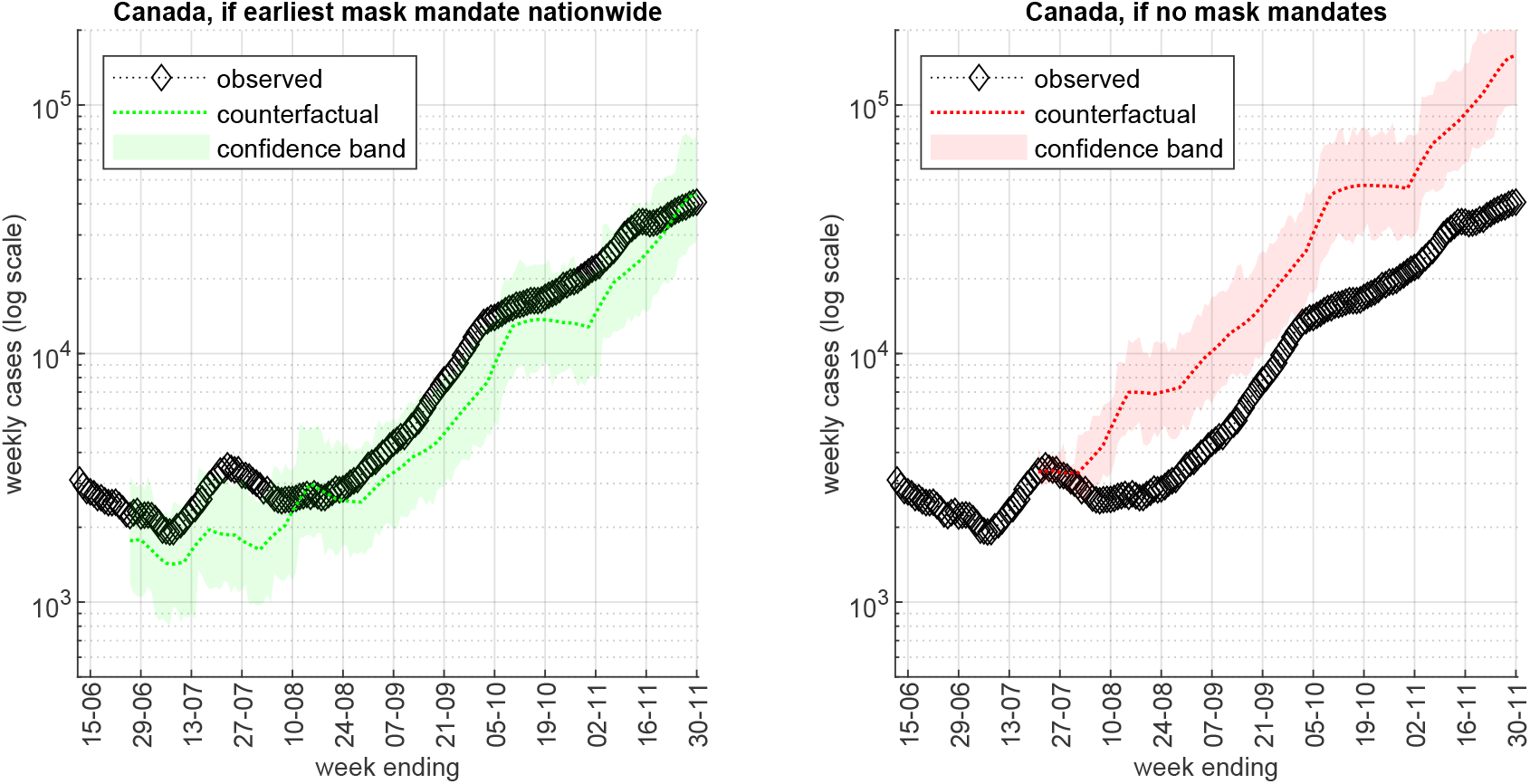
Canada – Counterfactuals Notes: The left panel assumes that mask mandates were adopted in all provinces on June 12, 2020 (date of the earliest mask mandate in Ontario). The right panel assumes that mask mandates were not adopted in any province. We use the estimates from column (2) of Table 2. The diamonds plot observed weekly cases from *t* −6 to *t*, the dotted lines plot the 7-day moving average of the counterfactual mean value, and the shaded areas are 5-95 percentile confidence bands.

In the right side panels of Fig. 8 and 9, we instead assume that mask mandates were *not* adopted in any Ontario PHU or in any Canadian province. Our estimates imply that such counterfactual absence of mask mandates, holding all else equal, would have led to a large and growing over time increase in new cases, both in Ontario (estimated 5,400 additional weekly cases at the end of September and 33,000 additional cases per week as of Nov. 30) and Canada-wide (estimated 10,700 additional weekly cases at the end of September and over 100,000 additional cases per week as of Nov. 30).

In Fig. C10 in the Appendix, we also simulate a counterfactual in which British Columbia (BC) and Alberta, the third and fourth largest Canadian provinces by population, adopt province-wide mask mandates on June 12. The results indicate an average reduction of about 1,000 cases per week for BC and 550 cases for Alberta over the simulation period. The estimated effect is smaller for Alberta, despite its larger case totals, since Alberta adopted mask mandates in its main cities on Aug. 1 while BC had no mask mandate until November.

The counterfactual simulations discussed above assume that behaviour, policies and all other variables (except mask policy and *t*−14 case counts) remain fixed at their values observed in the data. This is a strong assumption which may be more plausible over a relatively short period (e.g., focusing on July/August). In addition, all counterfactuals assume that regions without a mask mandate by a given date would react to a mandate in similar way, on average, as the regions that have already imposed a mandate. Therefore, these simulation results should be interpreted with caution and only as a rough illustration of the estimated impact of mask mandates on COVID-19 cases.

## 5 Conclusion

We estimate the impact of indoor face mask mandates and other public policy measures on the spread of COVID-19 in Canada. We use both within-province variation from Ontario and cross-province variation in the timing of mandates. We find robust and consistent significantly negative association between mask mandates and subsequent COVID-19 case growth – a 20 to 22% average weekly reduction in new cases. These results are supported by our analysis of survey data, which shows that the mandates led to a significant increase of 27 percentage points in the proportion of people reporting always wearing a mask in public. We show that the impact of mask mandates on COVID-19 case growth persists as long as they are necessary to achieve near universal mask wearing in indoor public spaces.

The earlier mask mandates in Canada were introduced in Summer 2020 when other policy measures were relaxed, as part of the economy’s re-opening. We find that reduced restrictions on businesses or gatherings are associated with subsequent COVID-19 case growth – a factor that can offset and obscure the public health benefits of mask mandates. Past case totals were also found to matter for subsequent COVID-19 outcomes, suggesting that riskier behaviour may follow prior information perceived as favourable. This feedback mechanism may limit how low the number of new cases can be pushed by mask mandates or other restrictions short of a lockdown. On the other hand, we find no evidence that mask mandates reduce adherence to other precautions against COVID-19. Indeed, they could potentially serve as an important behavioural anchor, especially in periods of plateauing or declining transmission.

Importantly, the effect of mask mandates that we estimate is *relative* to the absence of mandates and not absolute. In unfavourable conditions, such as during seasons when people gather indoors (fall, winter and early spring in most of Canada) and before widespread vaccination, a mask mandate appears insufficient to prevent an increase in new infections on its own and should be considered in conjunction with other policy measures.

We conclude that mandating mask wearing in indoor public spaces is a powerful policy tool to limit COVID-19 transmission, especially given its relatively low cost to the economy. As long as it remains doubtful that a sufficient fraction of the population has been immunized to achieve herd immunity – a concern particularly relevant in the face of more contagious COVID-19 variants and before vaccines are administered to children – it appears prudent to keep mask mandates in place even while lifting other restrictions.

We have deliberately refrained from studying direct economic impacts of COVID-19, focusing instead on unique features of the Canadian data for identifying the effect of mask mandates and other NPIs on COVID-19 case growth. Future research, jointly considering the epidemiological impact and the economic benefits and costs of the various public policies and restrictions, would enrich the ongoing debate and provide further guidance.

## Data Availability

All data used in this manuscript is based on information available in public domain, and is available via the data availability link and/or links provided in the manuscript.

https://github.com/C19-SFU-Econ/Public-Data

## Appendix A. Additional Analysis

### Closing period

We investigate whether the NPIs’ impact differed in the initial closing-down period of imposition of policy measures by restricting the sample to the period March 11 to May 14. The end date (referring to the NPIs in place on April 30) was chosen because very few NPIs were relaxed in Canada before early May (see Fig. 2). The results in Table B16 suggest that the imposition of closures, long-term care restrictions, and travel restrictions early in the pandemic was associated with a sharp subsequent reduction in weekly case growth, as also seen on Fig. C8 – the average log weekly growth rate of cases Δ log(ΔC) fell from 2.4 (ten-fold growth in weekly cases) to −0.4 (33% decrease in weekly cases) between March 15 and April 5. We interpret these results with caution, however, since many policy measures and restrictions were enacted in a short time interval during March 2020 and there is not much inter-provincial variation (see Fig. 2). No mask mandates were present in Canada in this period. We did not find any additional policy impacts in the mid-May to November period (not reported in the Table).

### Deaths

We also examine the weekly deaths growth rate as an outcome variable. Because COVID-19 deaths in Canada are highly concentrated in long-term care homes, mask mandates and NPIs may have a different effect on deaths growth than on cases. Table B17 reports estimates of equation (1) using the weekly deaths growth rate *Y*_*it*_ = Δ log(Δ*D*_*it*_) as the dependent variable, for each province *i* and date *t* in the sample period, where Δ*D*_*it*_ are weekly deaths from date *t*−6 to date *t*. We use a 28-day lag for the policy, behaviour, and information variables, to reflect the fact that deaths occur on average about two weeks after case detection (see Appendix E for detailed justification). The estimate coefficients on Mask 28 are not statistically significantly different from zero in our baseline specifications. However, considering the large standard errors, they are not inconsistent in magnitude with our estimates for mask mandates and case growth in Table 2. In addition, when weighing the data by provincial population in columns (7) and (8), which de-emphasizes the small provinces with very few deaths, the results in Table B17 suggest that mask mandates are associated with a 39 log points reduction in the weekly death rate (32% weekly reduction in deaths relative to the trend in absence of mandate). We supplement Table B17 with Fig. C11, where we display the estimated coefficients, with 95% confidence intervals, on the main variable of interest Mask 28 for different sample end dates between August 31 and November 30, 2020.

## Appendix B. Additional Tables

**Table B1:** Ontario – Correlations between the Google mobility indicators

| category | retail | grocery | workplaces | transit | residential | parks | N |
| --- | --- | --- | --- | --- | --- | --- | --- |
| retail | 1 |  |  |  |  |  | 6,672 |
| grocery | 0.78 | 1 |  |  |  |  | 6,735 |
| workplaces | 0.48 | 0.34 | 1 |  |  |  | 6,788 |
| transit | 0.55 | 0.44 | 0.62 | 1 |  |  | 5,040 |
| residential | -0.59 | -0.39 | -0.91 | -0.68 | 1 |  | 6,232 |
| parks | 0.32 | 0.23 | 0.23 | 0.39 | -0.34 | 1 | 3,014 |

Notes: Time period – May 1 to November 16, 2020 (two weeks before the May 15 - November 30 sample period). Daily PHU-level data.

**Table B2:** Canada – Correlations between the Google mobility indicators

| category | retail | grocery | workplaces | transit | residential | parks | N |
| --- | --- | --- | --- | --- | --- | --- | --- |
| retail | 1 |  |  |  |  |  | 2,650 |
| grocery | 0.81 | 1 |  |  |  |  | 2,650 |
| workplaces | 0.65 | 0.49 | 1 |  |  |  | 2,650 |
| transit | 0.77 | 0.53 | 0.81 | 1 |  |  | 2,306 |
| residential | -0.79 | -0.57 | -0.89 | -0.81 | 1 |  | 2,592 |
| parks | 0.50 | 0.41 | 0.15 | 0.25 | -0.46 | 1 | 2,168 |

Notes: Time period – February 26 to November 16, 2020 (two weeks before the March 11 - November 30 sample period). Daily province-level data.

**Table B3:** Ontario – Correlations between policies and location behaviour

| | Behaviour proxy | Mask | Business/<br>gathering | School | LTC | New weekly tests $\Delta T$ |
| --- | --- | --- | --- | --- | --- | --- |
| Behaviour proxy | 1 |  |  |  |  |  |
| Mask | 0.24 | 1 |  |  |  |  |
| Business/gathering | -0.50 | -0.70 | 1 |  |  |  |
| School | -0.43 | -0.62 | 0.89 | 1 |  |  |
| Long-term care (LTC) | -0.31 | -0.88 | 0.73 | 0.70 | 1 |  |
| New weekly tests $\Delta T$ | -0.44 | 0.19 | -0.02 | -0.08 | -0.14 | 1 |

Notes: Time period – May 15 to November 30, 2020 ( $N = 6,800$ ). Each variable is a 7-day moving average. All variables are at the PHU level, except School and LTC, which are measured at the province level.

**Table B4:** Canada – Correlations between policies and location behaviour

| | Behaviour proxy | Mask | Business/<br>gathering | School | Travel | LTC | New weekly tests $\Delta T$ |
| --- | --- | --- | --- | --- | --- | --- | --- |
| Behaviour proxy | 1 |  |  |  |  |  |  |
| Mask | 0.19 | 1 |  |  |  |  |  |
| Business/gathering | -0.84 | -0.33 | 1 |  |  |  |  |
| School | -0.35 | -0.53 | 0.37 | 1 |  |  |  |
| Travel | -0.05 | -0.06 | 0.22 | 0.22 | 1 |  |  |
| Long-term care (LTC) | -0.16 | -0.31 | 0.28 | 0.24 | 0.09 | 1 |  |
| New weekly tests $\Delta T$ | 0.11 | 0.51 | -0.02 | -0.24 | -0.32 | 0.10 | 1 |

Notes: Time period – March 11 to November 30, 2020 ( $N = 2,650$ ). Province-level, 7-day moving averages.

**Table B5:**
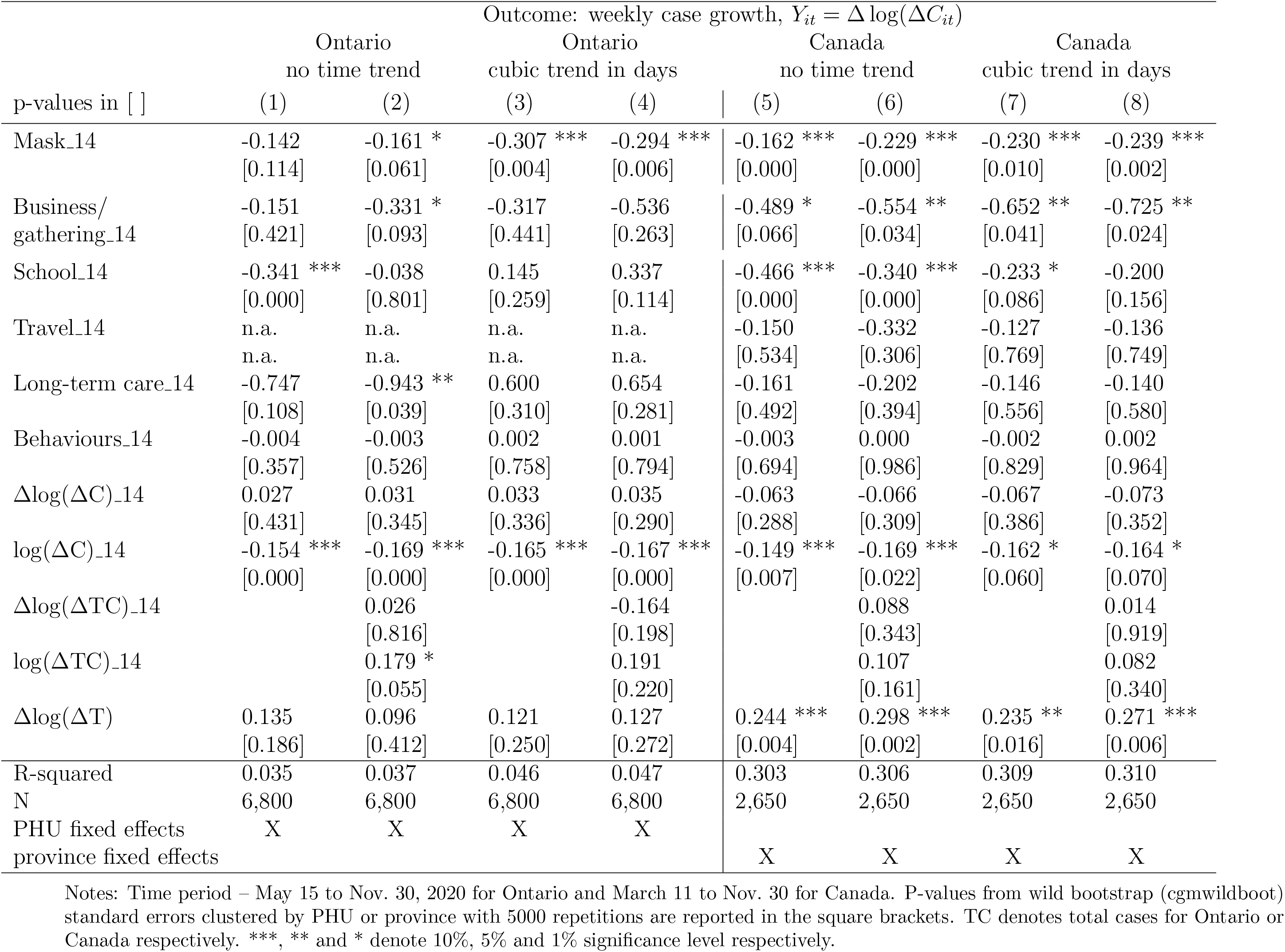
Alternative time controls

**Table B6:**
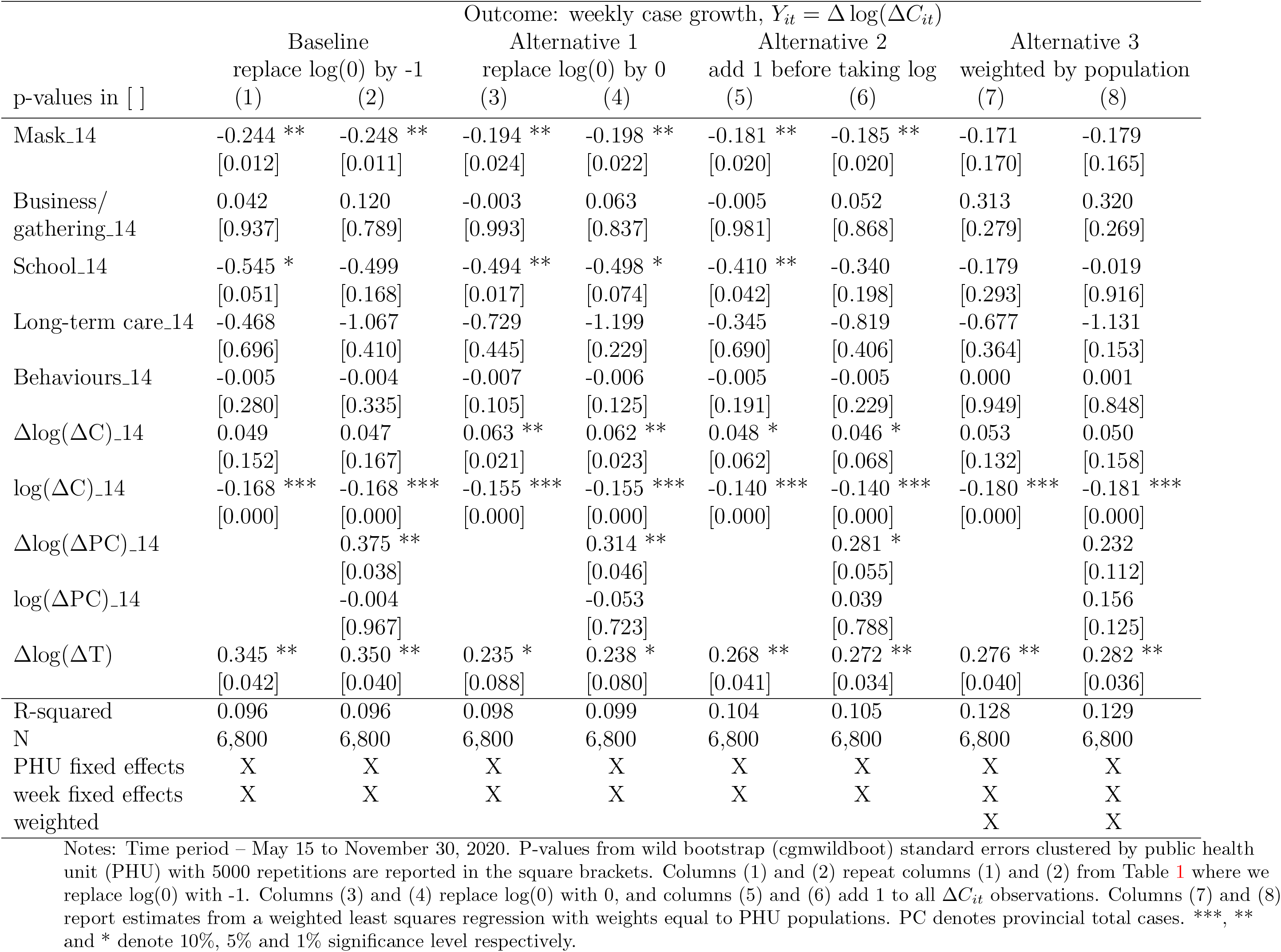
Ontario – Robustness (treatment of zero weekly cases)

**Table B7:** Ontario – Robustness (standard errors)

| Outcome: weekly case growth, $Y_{it} = \Delta \log(\Delta C_{it})$ | | | | |
| --- | --- | --- | --- | --- |
|  | (1) |  | (2) |  |
| Mask_14 | -0.244 |  | -0.248 |  |
|  | (0.017) | ** | (0.016) | ** |
|  | [0.012] | ** | [0.011] | ** |
|  | {0.023} | ** | {0.020} | ** |
| Business/gathering_14 | 0.042 |  | 0.120 |  |
|  | (0.921) |  | (0.777) |  |
|  | [0.937] |  | [0.789] |  |
|  | {0.945} |  | {0.814} |  |
| School_14 | -0.545 |  | -0.499 |  |
|  | (0.054) | * | (0.187) |  |
|  | [0.051] | * | [0.168] |  |
|  | {0.084} | * | {0.203} |  |
| Long-term care_14 | -0.468 |  | -1.067 |  |
|  | (0.710) |  | (0.405) |  |
|  | [0.696] |  | [0.410] |  |
|  | {0.690} |  | {0.422} |  |
| Behaviour proxy_14 | -0.005 |  | -0.004 |  |
|  | (0.288) |  | (0.335) |  |
|  | [0.280] |  | [0.335] |  |
|  | {0.301} |  | {0.327} |  |
| R-squared | 0.096 |  | 0.096 |  |
| N | 6,800 |  | 6,800 |  |
| $\Delta \log(\Delta C)_{14}$ | X | | X | |
| $\log(\Delta C)_{14}$ | X | | X | |
| $\Delta \log(\Delta PC)_{14}$ | | | X | |
| $\log(\Delta PC)_{14}$ | | | X | |
| $\Delta \log(\Delta T)$ | X | | X | |
| PHU fixed effects | X |  | X |  |
| week fixed effects | X |  | X |  |

Notes: Time period – May 15 to November 30. P-values from standard clustering by PHU (Stata command ‘cluster’) in the ( ) parentheses, wild bootstrap with one-way clustering by PHU and 5000 repetitions in the [ ] square brackets, and wild bootstrap with two-way clustering by PHU and day with 5000 repetitions in the { } curly braces. PC denotes provincial total cases. \*\*\*, \*\* and \* denote 10%, 5% and 1% significance level respectively.

**Table B8:**
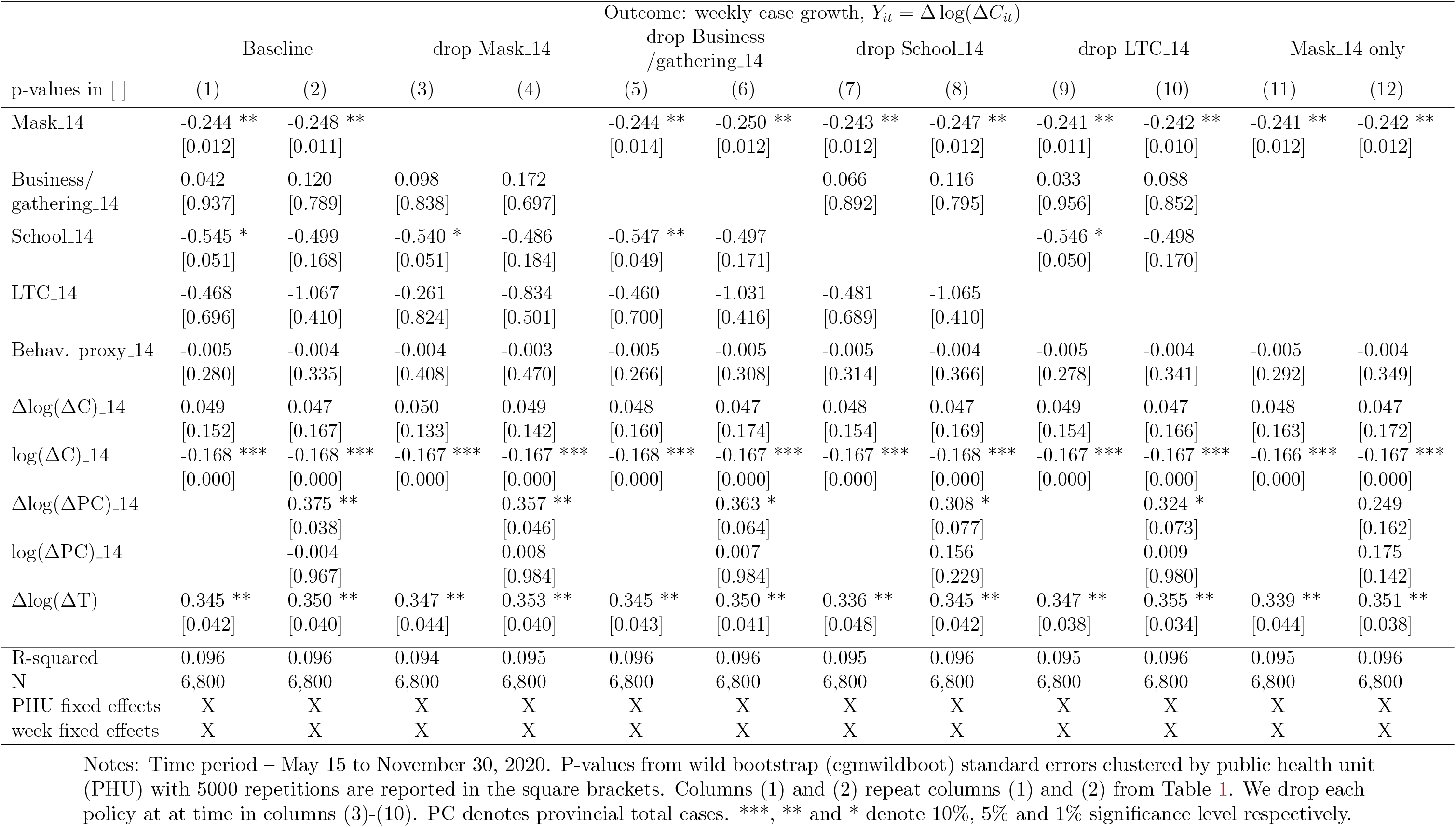
Ontario – Robustness (policy collinearity)

**Table B9:**
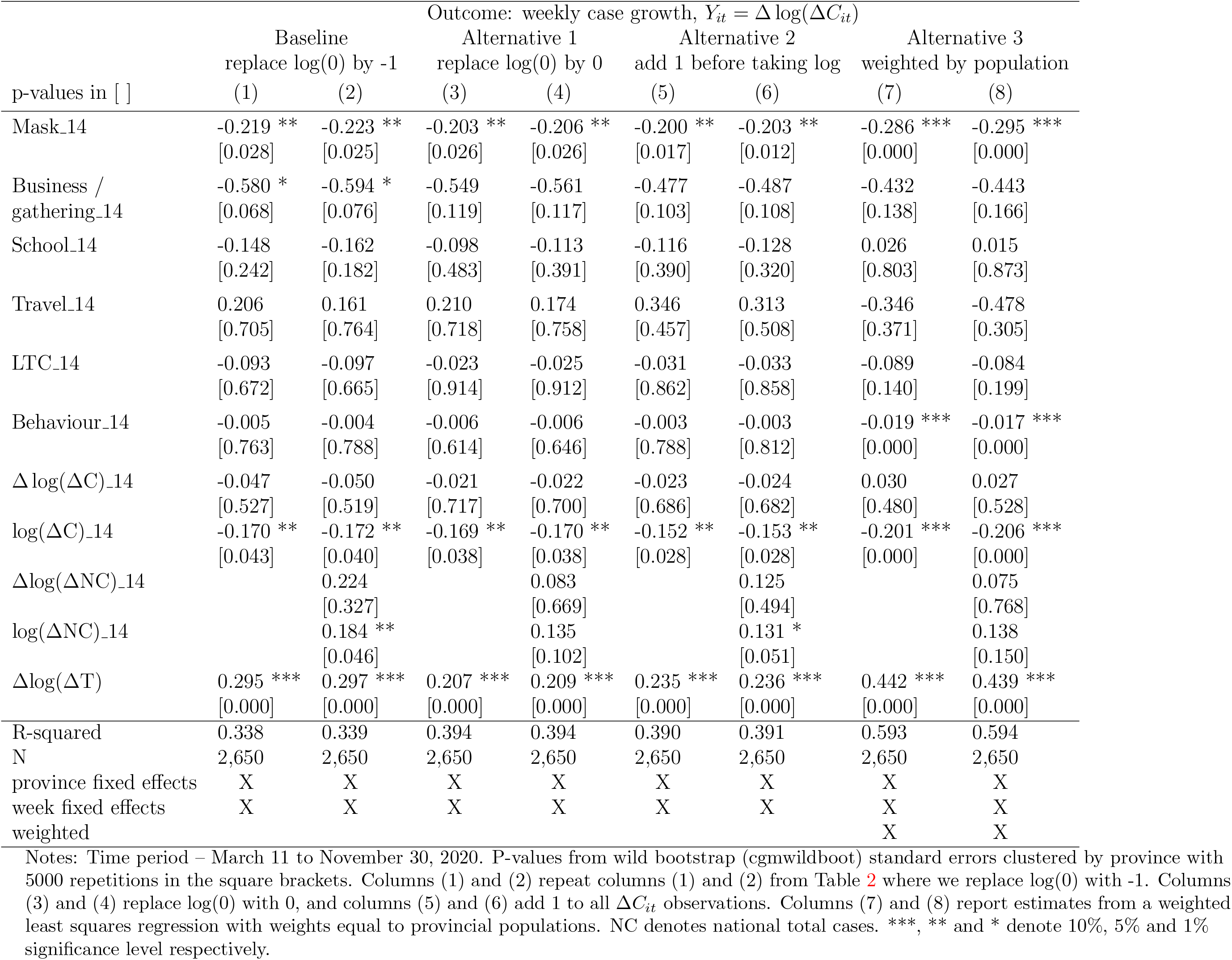
Canada - Robustness (treatment of zero weekly cases)

**Table B10:** Canada – Robustness (standard errors)

| Outcome: weekly case growth, $Y_{it} = \Delta \log(\Delta C_{it})$ | | | |
| --- | --- | --- | --- |
|  | (1) | (2) |  |
| Mask_14 | -0.219<br>(0.034) **<br>[0.028] **<br>{0.024} ** | -0.223<br>(0.032) **<br>[0.025] **<br>{0.017} ** |  |
| Business/gathering_14 | -0.580<br>(0.063) *<br>[0.068] *<br>{0.060} * | -0.594<br>(0.065) *<br>[0.076] *<br>{0.056} * |  |
| School_14 | -0.148<br>(0.301)<br>[0.242]<br>{0.210} | -0.162<br>(0.252)<br>[0.182]<br>{0.152} |  |
| Travel_14 | 0.206<br>(0.660)<br>[0.705]<br>{0.714} | 0.161<br>(0.733)<br>[0.764]<br>{0.761} |  |
| Long-term care_14 | -0.093<br>(0.655)<br>[0.672]<br>{0.672} | -0.097<br>(0.647)<br>[0.665]<br>{0.627} |  |
| Behaviour proxy_14 | -0.005<br>(0.713)<br>[0.763]<br>{0.760} | -0.004<br>(0.738)<br>[0.788]<br>{0.782} |  |
| R-squared | 0.338 | 0.339 |  |
| N | 2,650 | 2,650 |  |
| $\Delta \log(\Delta C)_{14}$ | X | X | |
| $\log(\Delta C)_{14}$ | X | X | |
| $\Delta \log(\Delta NC)_{14}$ | | X | |
| $\log(\Delta NC)_{14}$ | | X | |
| $\Delta \log(\Delta T)$ | X | X | |
| province fixed effects | X | X |  |
| week fixed effects | X | X |  |

Notes: Time period – March 11 to November 30. P-values from standard clustering by province in the ( ) parentheses, wild bootstrap with one-way clustering by province and 5000 repetitions in the [ ] square brackets, and wild bootstrap with two-way clustering by province and day with 5000 repetitions in the { } curly braces. NC denotes national total cases. \*\*\*, \*\* and \* denote 10%, 5% and 1% significance level respectively.

**Table B11:**
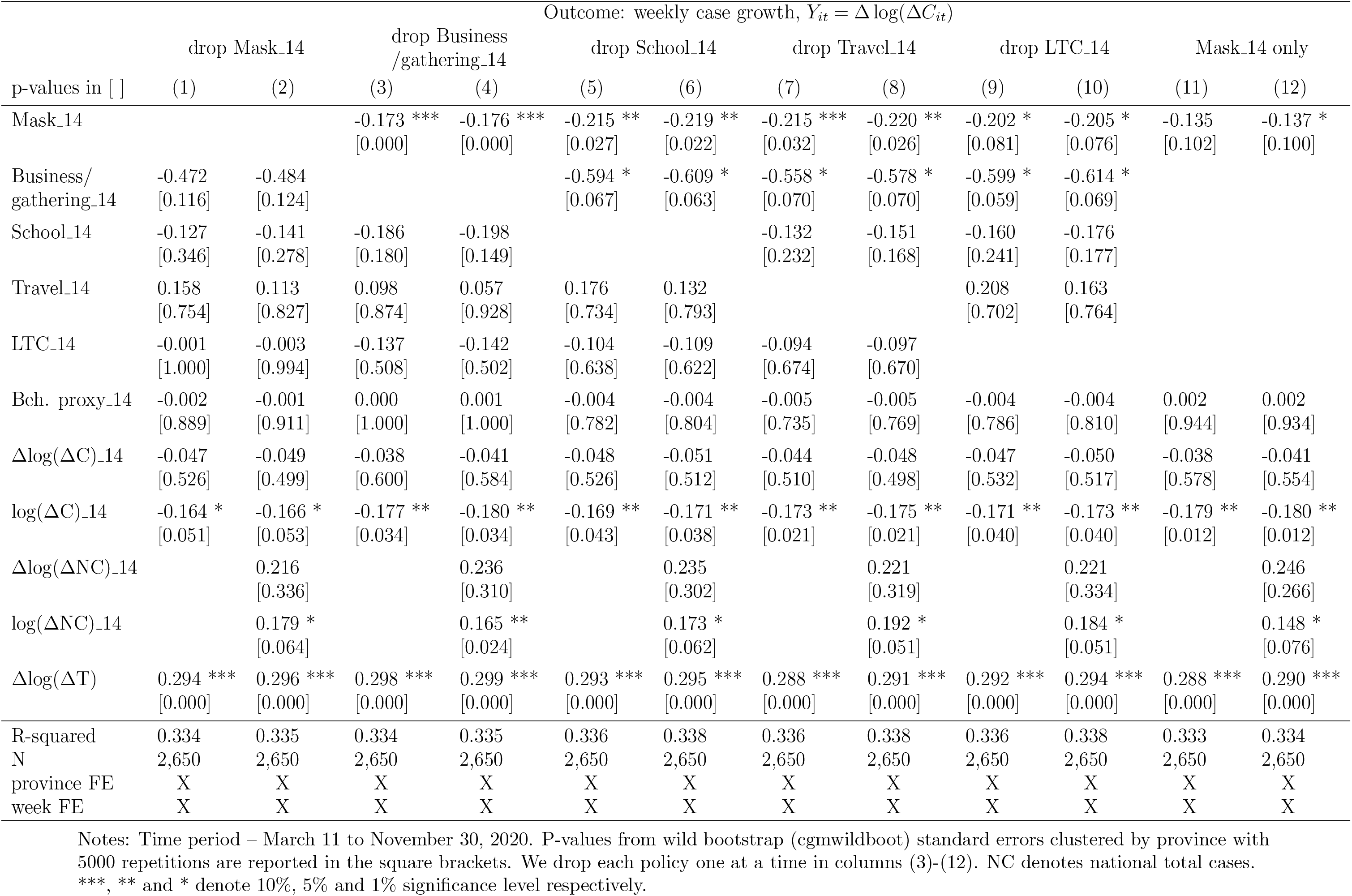
Canada - Robustness (policy collinearity)

**Table B12:** Canada – Robustness (news and weather)

| p-values in [ ] | Outcome: weekly case growth, $Y_{it} = \Delta \log(\Delta C_{it})$ | | | | | |
| --- | --- | --- | --- | --- | --- | --- |
|  | baseline |  | add weather |  | add news |  |
|  | (1) | (2) | (3) | (4) | (5) | (6) |
| Mask_14 | -0.219 **<br>[0.028] | -0.223 **<br>[0.025] | -0.186 **<br>[0.028] | -0.189 **<br>[0.025] | -0.217 **<br>[0.036] | -0.217 **<br>[0.029] |
| Business/gathering_14 | -0.580 *<br>[0.068] | -0.594 *<br>[0.076] | -0.605 *<br>[0.095] | -0.623 *<br>[0.090] | -0.574 *<br>[0.075] | -0.573 *<br>[0.076] |
| School_14 | -0.148<br>[0.242] | -0.162<br>[0.182] | -0.105<br>[0.503] | -0.120<br>[0.416] | -0.140<br>[0.256] | -0.126<br>[0.294] |
| Travel_14 | 0.206<br>[0.705] | 0.161<br>[0.764] | 0.149<br>[0.773] | 0.091<br>[0.846] | 0.217<br>[0.693] | 0.184<br>[0.732] |
| Long-term care_14 | -0.093<br>[0.672] | -0.097<br>[0.665] | -0.084<br>[0.711] | -0.087<br>[0.711] | -0.091<br>[0.684] | -0.089<br>[0.690] |
| Behaviour proxy_14 | -0.005<br>[0.763] | -0.004<br>[0.788] | -0.003<br>[0.807] | -0.003<br>[0.842] | -0.005<br>[0.769] | -0.002<br>[0.873] |
| $\Delta \log(\Delta C)$_14 | -0.047<br>[0.527] | -0.050<br>[0.519] | -0.048<br>[0.497] | -0.051<br>[0.487] | -0.047<br>[0.527] | -0.052<br>[0.500] |
| $\log(\Delta C)$_14 | -0.170 **<br>[0.043] | -0.172 **<br>[0.040] | -0.171 **<br>[0.035] | -0.174 **<br>[0.031] | -0.169 **<br>[0.048] | -0.168 **<br>[0.045] |
| $\Delta \log(\Delta NC)$_14 | | 0.224<br>[0.327] | | 0.207<br>[0.356] | | 0.381<br>[0.109] |
| $\log(\Delta NC)$_14 | | 0.184 **<br>[0.046] | | 0.209 **<br>[0.020] | | 0.412 ***<br>[0.009] |
| $\Delta \log(\Delta T)$ | 0.295 ***<br>[0.000] | 0.297 ***<br>[0.000] | 0.316 ***<br>[0.000] | 0.318 ***<br>[0.000] | 0.295 ***<br>[0.000] | 0.293 ***<br>[0.000] |
| Rain_14 |  |  | 0.019<br>[0.461] | 0.019<br>[0.478] |  |  |
| Max temp_14 |  |  | 0.034<br>[0.333] | 0.035<br>[0.336] |  |  |
| Min temp_14 |  |  | -0.043<br>[0.270] | -0.045<br>[0.260] |  |  |
| News_14 |  |  |  |  | -0.002<br>[0.680] | -0.014 *<br>[0.068] |
| R-squared | 0.338 | 0.339 | 0.340 | 0.342 | 0.337 | 0.340 |
| N | 2,650 | 2,650 | 2,650 | 2,650 | 2,650 | 2,650 |
| province fixed effects | X | X | X | X | X | X |
| week fixed effects | X | X | X | X | X | X |
| weather |  |  | X | X |  |  |
| news |  |  |  |  | X | X |

Notes: Time period – March 11 to November 30, 2020. P-values from wild bootstrap (cgmwildboot) standard errors clustered by province with 5000 repetitions are reported in the square brackets. Columns (1) and (2) repeat columns (1) and (2) from Table 2. Columns (3) and (4) report estimates with lagged weather variables as additional controls. Columns (5) and (6) add a “news” variable to the baseline specification (see Appendix D for more details). NC denotes national total cases. \*\*\*, \*\* and \* denote 10%, 5% and 1% significance level respectively.

**Table B13:**
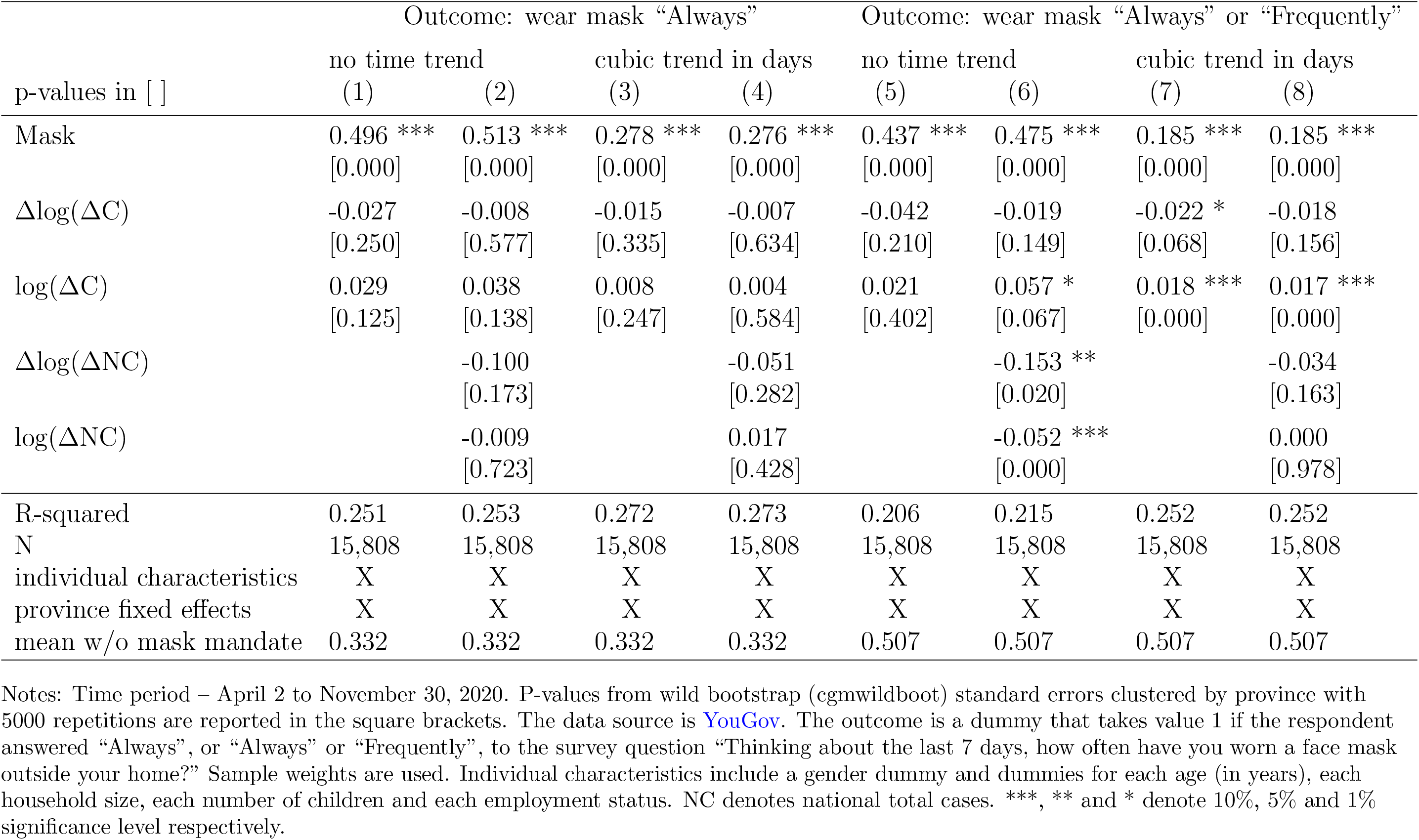
Self-reported mask usage – Alternative time controls

**Table B14:** Self-reported precautions – Canada

| p-values in [ ] | Outcome: “Always” response |  |  |  |  |  |  |  |
| --- | --- | --- | --- | --- | --- | --- | --- | --- |
|  | wash hands | use sanitizer | avoid going out in general | avoid small gatherings | avoid medium gatherings | avoid large gatherings | avoid crowded areas | avoid touching objects |
|  | (1) | (2) | (3) | (4) | (5) | (6) | (7) | (8) |
| Mask | 0.001<br>[0.955] | -0.001<br>[0.972] | 0.004<br>[0.636] | 0.046 *<br>[0.056] | 0.043<br>[0.238] | 0.065<br>[0.118] | 0.047<br>[0.174] | 0.027<br>[0.340] |
| $\Delta\log(\Delta C)$ | -0.001<br>[0.368] | -0.003<br>[0.622] | -0.005<br>[0.340] | -0.007<br>[0.386] | -0.006<br>[0.548] | -0.006<br>[0.529] | 0.005<br>[0.645] | 0.001<br>[0.905] |
| $\log(\Delta C)$ | -0.004<br>[0.421] | 0.002<br>[0.567] | 0.019 ***<br>[0.000] | 0.024 ***<br>[0.000] | 0.025 ***<br>[0.005] | 0.022 **<br>[0.029] | 0.011<br>[0.160] | 0.007 *<br>[0.076] |
| $\Delta\log(\Delta NC)$ | -0.108<br>[0.202] | 0.080<br>[0.528] | 0.026<br>[0.664] | 0.053<br>[0.389] | 0.103<br>[0.279] | 0.080<br>[0.209] | 0.122 ***<br>[0.000] | 0.020<br>[0.731] |
| $\log(\Delta NC)$ | 0.125<br>[0.101] | 0.108<br>[0.246] | -0.037<br>[0.340] | 0.067<br>[0.363] | 0.064<br>[0.394] | 0.109<br>[0.200] | 0.009<br>[0.788] | 0.056<br>[0.204] |
| R-squared | 0.037 | 0.049 | 0.039 | 0.101 | 0.112 | 0.103 | 0.069 | 0.034 |
| N | 15,808 | 15,808 | 15,808 | 15,808 | 15,808 | 15,808 | 15,808 | 15,808 |
| individual characteristics | X | X | X | X | X | X | X | X |
| province fixed effects | X | X | X | X | X | X | X | X |
| week fixed effects | X | X | X | X | X | X | X | X |
| average w/o mask mandate | 0.721 | 0.489 | 0.259 | 0.444 | 0.575 | 0.755 | 0.640 | 0.476 |
| survey item # (i12_) | health_2 | health_3 | health_6 | health_12 | health_13 | health_14 | health_15 | health_20 |

Notes: Time period – April 2 to November 30, 2020. P-values from wild bootstrap (cgwildboot) standard errors clustered by province with 5000 repetitions are reported in the square brackets. The data source is YouGov. The outcome is a dummy that takes value 1 if the respondent answered “Always” to each survey question listed in Table D5. Sample weights are used. Individual characteristics include a gender dummy and dummies for each age (in years), each household size, each number of children and each employment status. NC denotes national total cases. \*\*\*, \*\* and \* denote 10%, 5% and 1% significance level respectively.

**Table B15:**
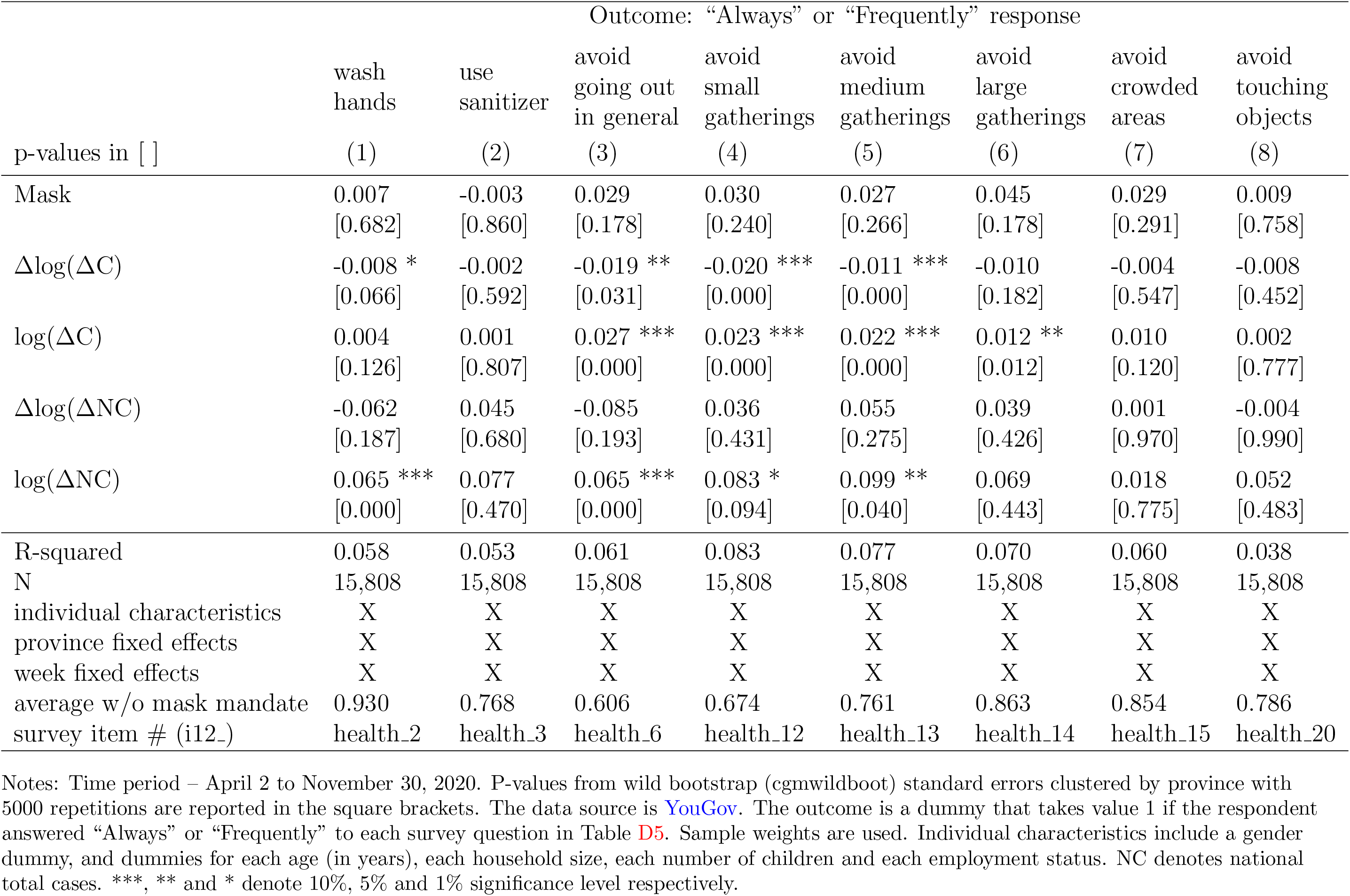
Self-reported precautions – Canada (continued)

**Table B16:** Canada – Initial closing sub-period

| Outcome: weekly case growth, $Y_{it} = \Delta \log(\Delta C_{it})$ | | | | |
| --- | --- | --- | --- | --- |
|  | Baseline |  | Initial closing |  |
|  | Mar. 11 - Nov. 30 |  | Mar. 11 - May 14 |  |
| p-values in [ ] | (1) | (2) | (3) | (4) |
| Mask_14 | -0.219 **<br>[0.028] | -0.223 **<br>[0.025] | n.a.<br>n.a. | n.a.<br>n.a. |
| Business/gathering_14 | -0.580 *<br>[0.068] | -0.594 *<br>[0.076] | -0.141<br>[0.852] | -0.227<br>[0.775] |
| School_14 | -0.148<br>[0.242] | -0.162<br>[0.182] | -0.543<br>[0.221] | -0.698<br>[0.160] |
| Travel_14 | 0.206<br>[0.705] | 0.161<br>[0.764] | -1.876 ***<br>[0.000] | -2.315 ***<br>[0.000] |
| Long-term care_14 | -0.093<br>[0.672] | -0.097<br>[0.665] | -0.647 ***<br>[0.000] | -0.864 ***<br>[0.000] |
| Behaviour proxy_14 | -0.005<br>[0.763] | -0.004<br>[0.788] | -0.031 **<br>[0.010] | -0.031 **<br>[0.040] |
| $\Delta \log(\Delta C)_{14}$ | -0.047<br>[0.527] | -0.050<br>[0.519] | 0.075<br>[0.295] | 0.074<br>[0.268] |
| $\log(\Delta C)_{14}$ | -0.170 **<br>[0.043] | -0.172 **<br>[0.040] | -0.350 ***<br>[0.000] | -0.394 ***<br>[0.000] |
| $\Delta \log(\Delta NC)_{14}$ | | 0.224<br>[0.327] | | 0.238<br>[0.347] |
| $\log(\Delta NC)_{14}$ | | 0.184 **<br>[0.046] | | 0.457 ***<br>[0.000] |
| $\Delta \log(\Delta T)$ | 0.295 ***<br>[0.000] | 0.297 ***<br>[0.000] | 0.198 **<br>[0.019] | 0.197 **<br>[0.031] |
| R-squared | 0.338 | 0.339 | 0.673 | 0.684 |
| N | 2,650 | 2,650 | 650 | 650 |
| province fixed effects | X | X | X | X |
| week fixed effects | X | X | X | X |

Notes: P-values from wild bootstrap (cgmwildboot) standard errors clustered by province with 5000 repetitions are reported in the square brackets. NC denotes national total cases. \*\*\*, \*\* and \* denote 10%, 5% and 1% significance level respectively. No mask mandates are present in the initial closing period.

**Table B17:**
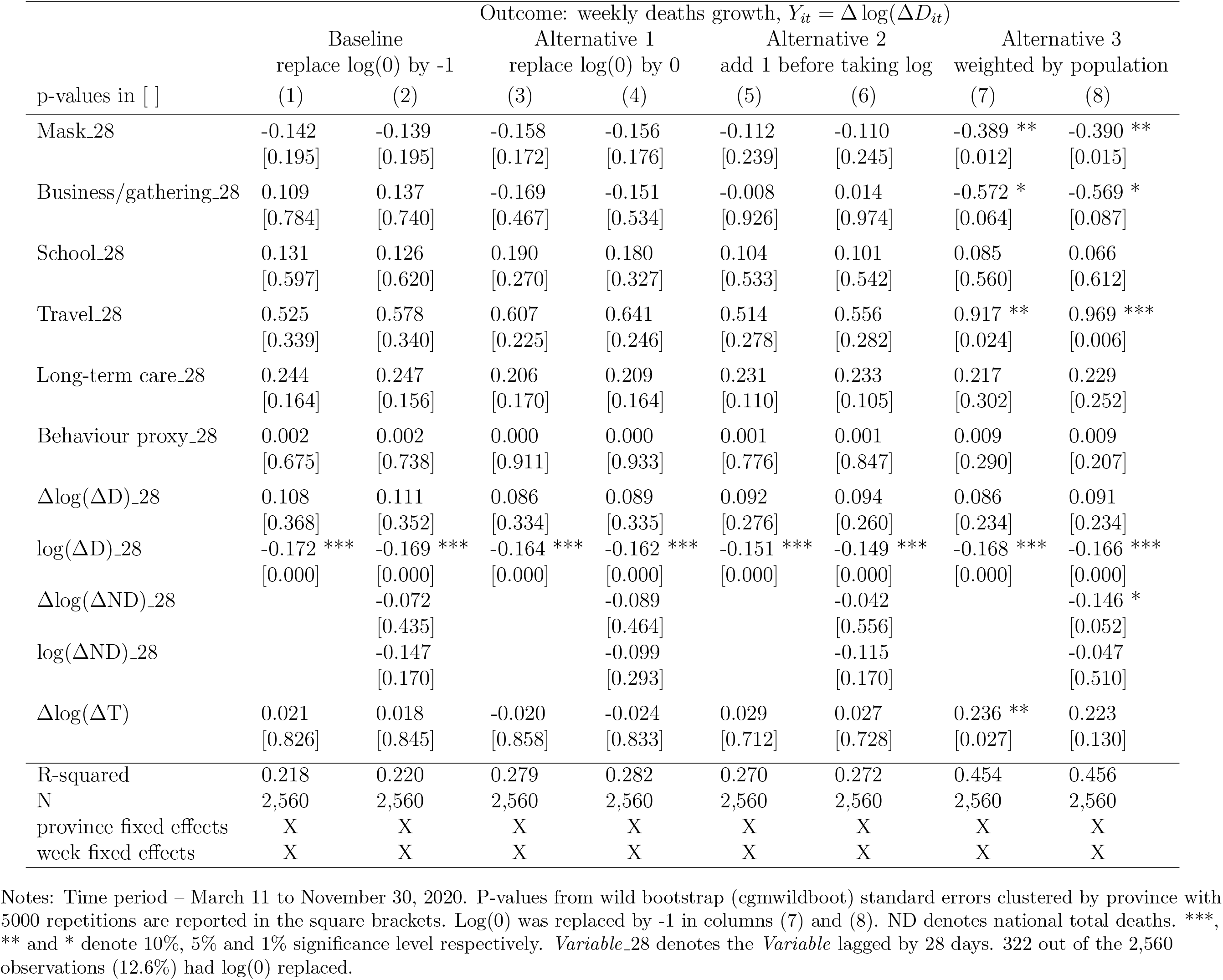
Canada – deaths growth rate

**Table B18:**
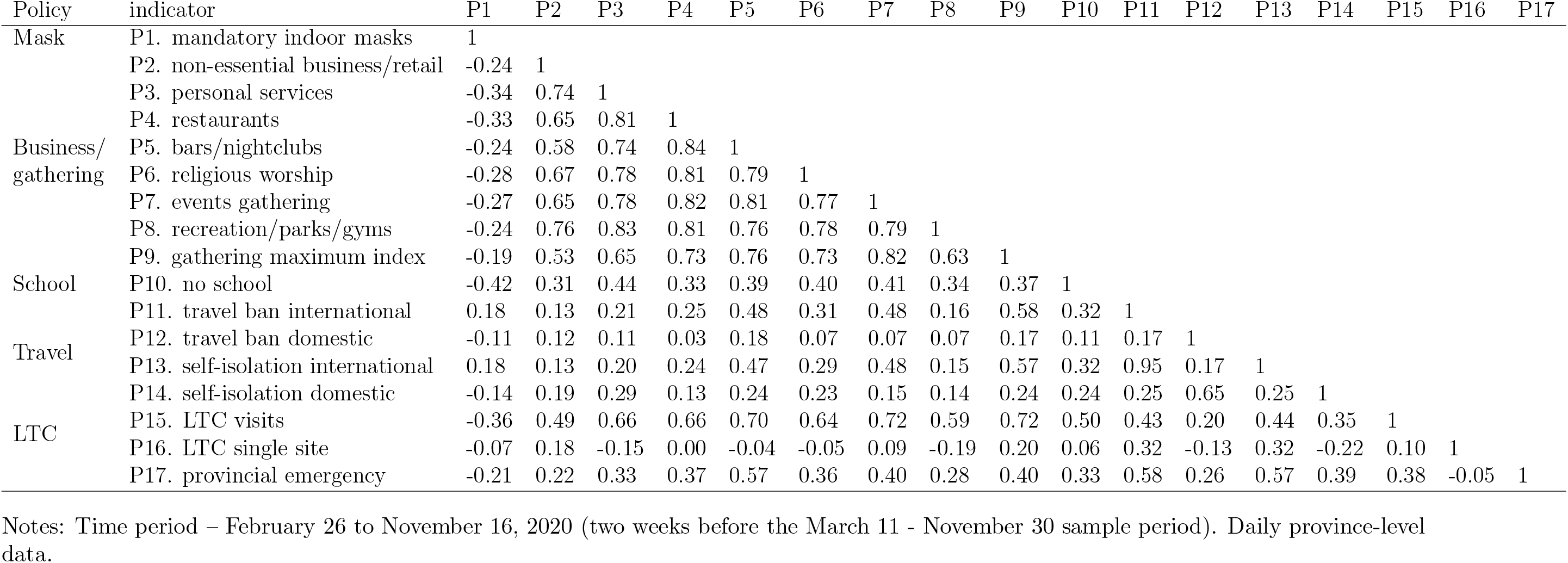
Canada - Correlations between the policy indicators

**Table B19:** Canada – Location behaviour and policies

| p-values in [ ] | Outcome: location behaviour |  |  |  |  |  |  |  |
| --- | --- | --- | --- | --- | --- | --- | --- | --- |
| | Retail | | Grocery | | Workplaces | | Behaviour proxy, $B_{it}$ | |
|  | (1) | (2) | (3) | (4) | (5) | (6) | (7) | (8) |
| Mask | -4.166 *<br>[0.077] | -4.019<br>[0.111] | -2.603<br>[0.178] | -2.558<br>[0.170] | -1.772 **<br>[0.018] | -1.631 **<br>[0.042] | -2.847 *<br>[0.064] | -2.736 *<br>[0.060] |
| Business/gathering | -21.18 ***<br>[0.004] | -20.14 **<br>[0.013] | -6.515<br>[0.185] | -5.672<br>[0.282] | -13.12 ***<br>[0.000] | -12.04 ***<br>[0.007] | -13.60 **<br>[0.016] | -12.62 **<br>[0.028] |
| School | 0.416<br>[0.859] | 1.599<br>[0.477] | -0.663<br>[0.756] | 0.188<br>[0.914] | -6.268 ***<br>[0.000] | -5.058 ***<br>[0.000] | -2.172<br>[0.265] | -1.090<br>[0.538] |
| Travel | -9.850<br>[0.549] | -7.243<br>[0.633] | -7.403<br>[0.234] | -5.837<br>[0.334] | -2.080<br>[0.740] | 0.537<br>[0.916] | -6.445<br>[0.441] | -4.181<br>[0.620] |
| Long-term care | -0.196<br>[0.951] | -0.065<br>[0.971] | -3.163<br>[0.118] | -3.189 *<br>[0.080] | -4.448<br>[0.138] | -4.332<br>[0.146] | -2.602<br>[0.326] | -2.529<br>[0.381] |
| $\Delta \log(\Delta C)$ | 1.121 *<br>[0.064] | 1.093 *<br>[0.068] | 0.950 **<br>[0.022] | 0.831 **<br>[0.016] | 0.457<br>[0.197] | 0.413<br>[0.213] | 0.843 ***<br>[0.000] | 0.779 ***<br>[0.006] |
| $\log(\Delta C)$ | -1.780 ***<br>[0.002] | -1.729 ***<br>[0.005] | -1.215 ***<br>[0.004] | -1.220 ***<br>[0.004] | -0.794 ***<br>[0.000] | -0.748 ***<br>[0.000] | -1.263 ***<br>[0.006] | -1.232 ***<br>[0.006] |
| $\Delta \log(\Delta NC)$ | | 3.813 *<br>[0.080] | | 11.69 ***<br>[0.000] | | 5.293 ***<br>[0.003] | | 6.933 ***<br>[0.000] |
| $\log(\Delta NC)$ | | -6.293 **<br>[0.016] | | -1.963<br>[0.225] | | -6.035 ***<br>[0.000] | | -4.764 ***<br>[0.007] |
| R-squared | 0.943 | 0.945 | 0.901 | 0.908 | 0.954 | 0.957 | 0.947 | 0.950 |
| N | 2,650 | 2,650 | 2,650 | 2,650 | 2,650 | 2,650 | 2,650 | 2,650 |
| province fixed effects | X | X | X | X | X | X | X | X |
| week fixed effects | X | X | X | X | X | X | X | X |

Notes: Time period – March 11 to November 30, 2020. P-values from wild bootstrap (cgmwildboot) standard errors clustered by province with 5000 repetitions are reported in the square brackets. The behaviour proxy $B_{it}$ in columns (7) and (8) is the arithmetic average of the “retail”, “grocery” and “workplaces” Google mobility indices. NC denotes national total cases. \*\*\*, \*\* and \* denote 10%, 5% and 1% significance level respectively.

**Table B20:** Policies and information only

| Outcome: weekly case growth, $Y_{it} = \Delta \log(\Delta C'_{it})$ | | | | |
| --- | --- | --- | --- | --- |
| p-values in [ ] | Ontario |  | Canada |  |
|  | (1) | (2) | (3) | (4) |
| Mask_14 | -0.234 **<br>[0.014] | -0.240 **<br>[0.013] | -0.204 ***<br>[0.000] | -0.210 ***<br>[0.000] |
| Business/gathering_14 | 0.084<br>[0.862] | 0.158<br>[0.721] | -0.525 *<br>[0.088] | -0.548 *<br>[0.096] |
| School_14 | -0.529 *<br>[0.058] | -0.475<br>[0.200] | -0.134<br>[0.309] | -0.152<br>[0.229] |
| Travel_14 | n.a.<br>n.a. | n.a.<br>n.a. | 0.227<br>[0.646] | 0.175<br>[0.733] |
| Long-term care_14 | -0.439<br>[0.706] | -1.052<br>[0.415] | -0.082<br>[0.742] | -0.087<br>[0.742] |
| $\Delta \log(\Delta C)_{14}$ | 0.049<br>[0.156] | 0.047<br>[0.171] | -0.052<br>[0.432] | -0.054<br>[0.422] |
| $\log(\Delta C)_{14}$ | -0.167 ***<br>[0.000] | -0.167 ***<br>[0.000] | -0.163 **<br>[0.028] | -0.166 **<br>[0.028] |
| $\Delta \log(\Delta TC)_{14}$ | | 0.379 **<br>[0.038] | | 0.207<br>[0.396] |
| $\log(\Delta TC)_{14}$ | | 0.007<br>[0.982] | | 0.197 ***<br>[0.008] |
| $\Delta \log(\Delta T)$ | 0.339 **<br>[0.047] | 0.345 **<br>[0.044] | 0.296 ***<br>[0.000] | 0.298 ***<br>[0.000] |
| R-squared | 0.095 | 0.096 | 0.336 | 0.338 |
| N | 6,800 | 6,800 | 2,650 | 2,650 |
| PHU fixed effects | X | X |  |  |
| province fixed effects |  |  | X | X |
| week fixed effects | X | X | X | X |

Notes: Time period – May 15 to November 30, 2020 for Ontario and March 11 to November 30, 2020 for Canada. P-values from wild bootstrap (cgmwildboot) standard errors clustered by PHU or province with 5000 repetitions are reported in the square brackets. TC denotes total cases for Ontario or Canada respectively. \*\*\*, \*\* and \* denote 10%, 5% and 1% significance level respectively..

## Appendix C. Additional Figures

**Figure C1:**
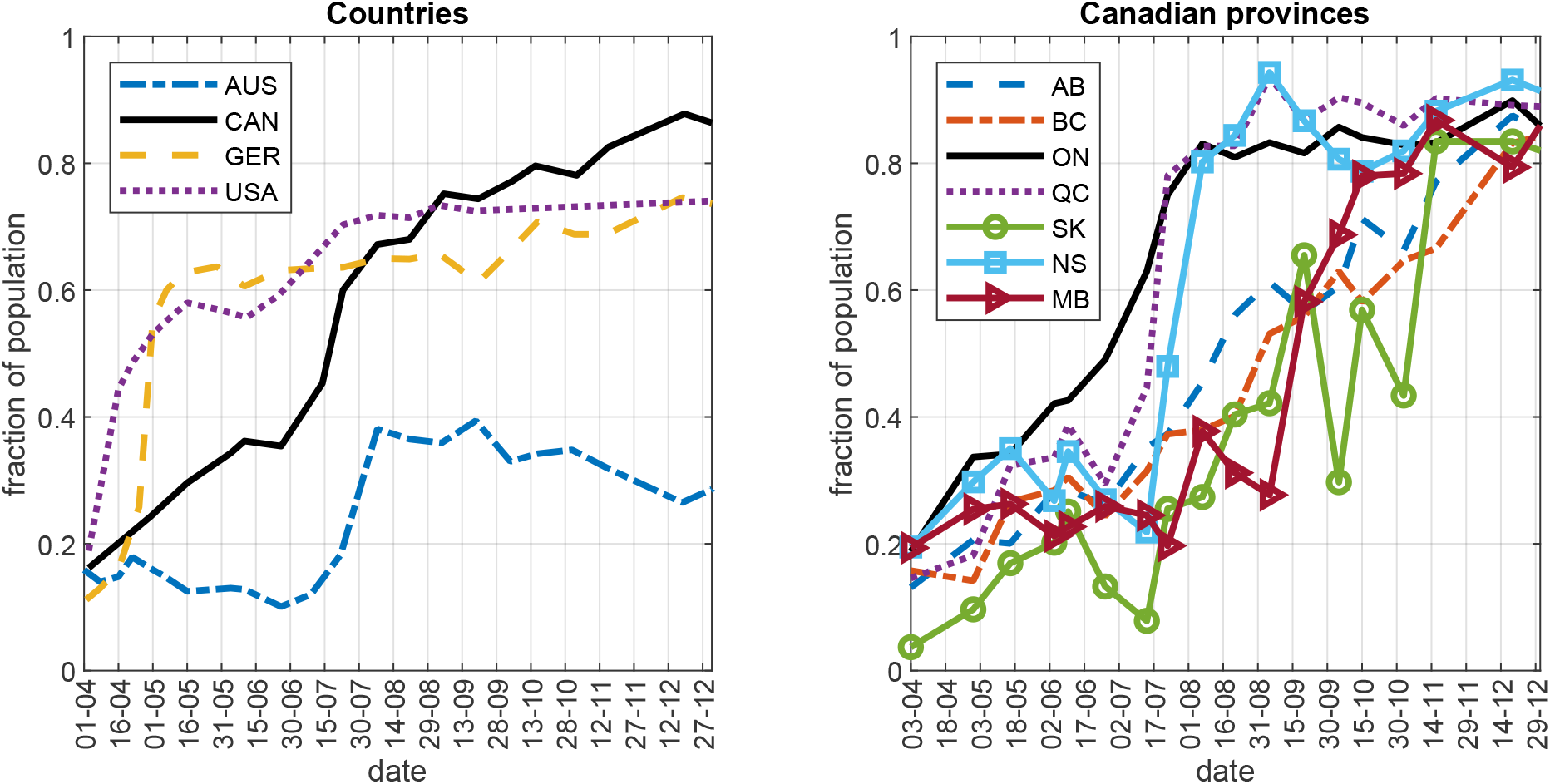
Self-reported mask usage in selected countries and Canadian provinces Notes: The data source is YouGov. The figure plots the average self-reported mask usage by week (the fraction of respondents that answered “Always” to the survey question “Thinking about the last 7 days, how often have you worn a face mask outside your home?”). Sample weights are used to compute the national and provincial averages.

**Figure C2:**
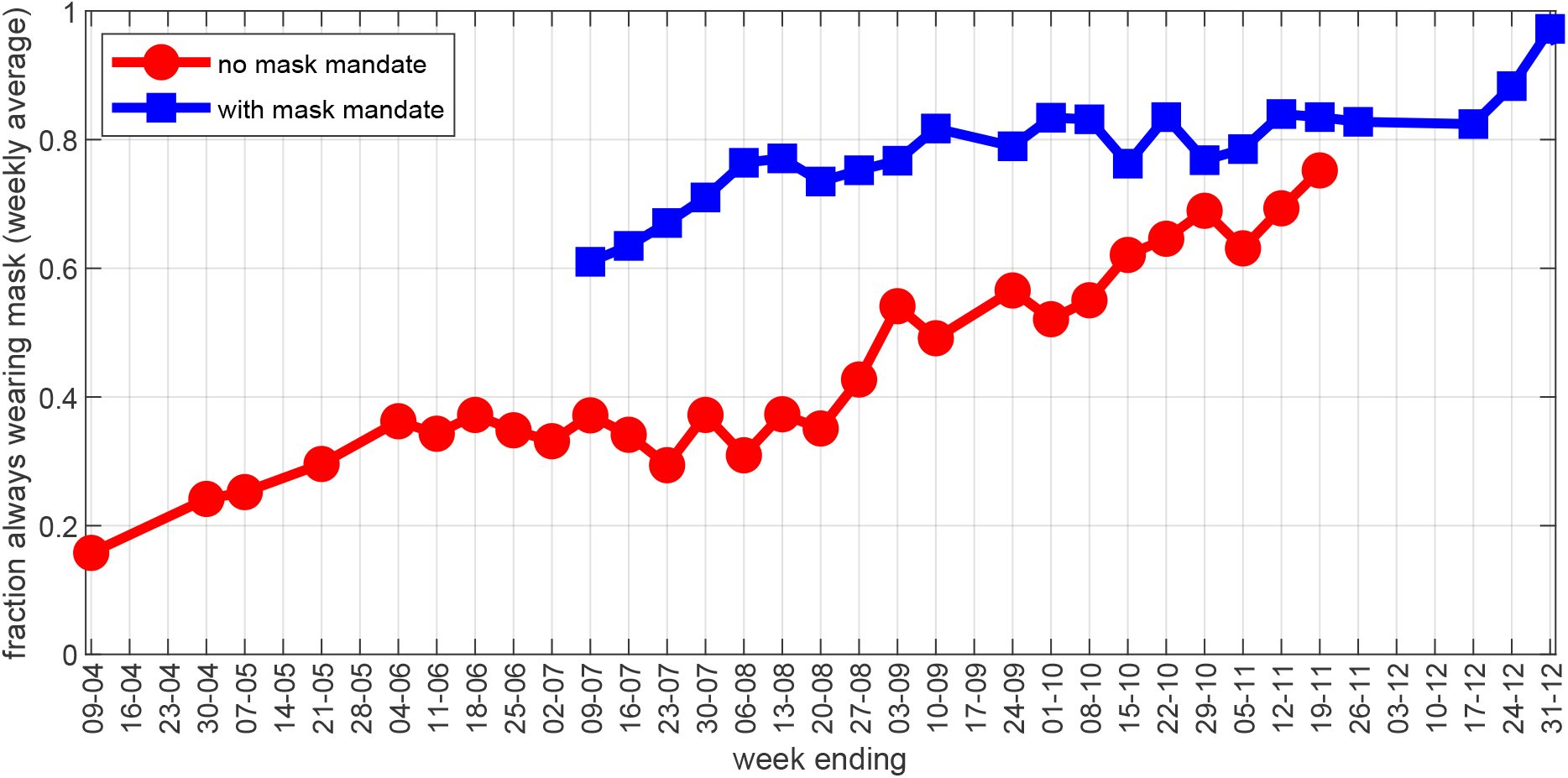
Canada – mask mandates and self-reported mask usage Notes: The data source is YouGov. The figure plots the average self-reported mask usage by week (the fraction of respondents that answered “Always” to the survey question “Thinking about the last 7 days, how often have you worn a face mask outside your home?”) in the provinces with and without mask mandates. Sample weights are used to compute the averages.

**Figure C3:**
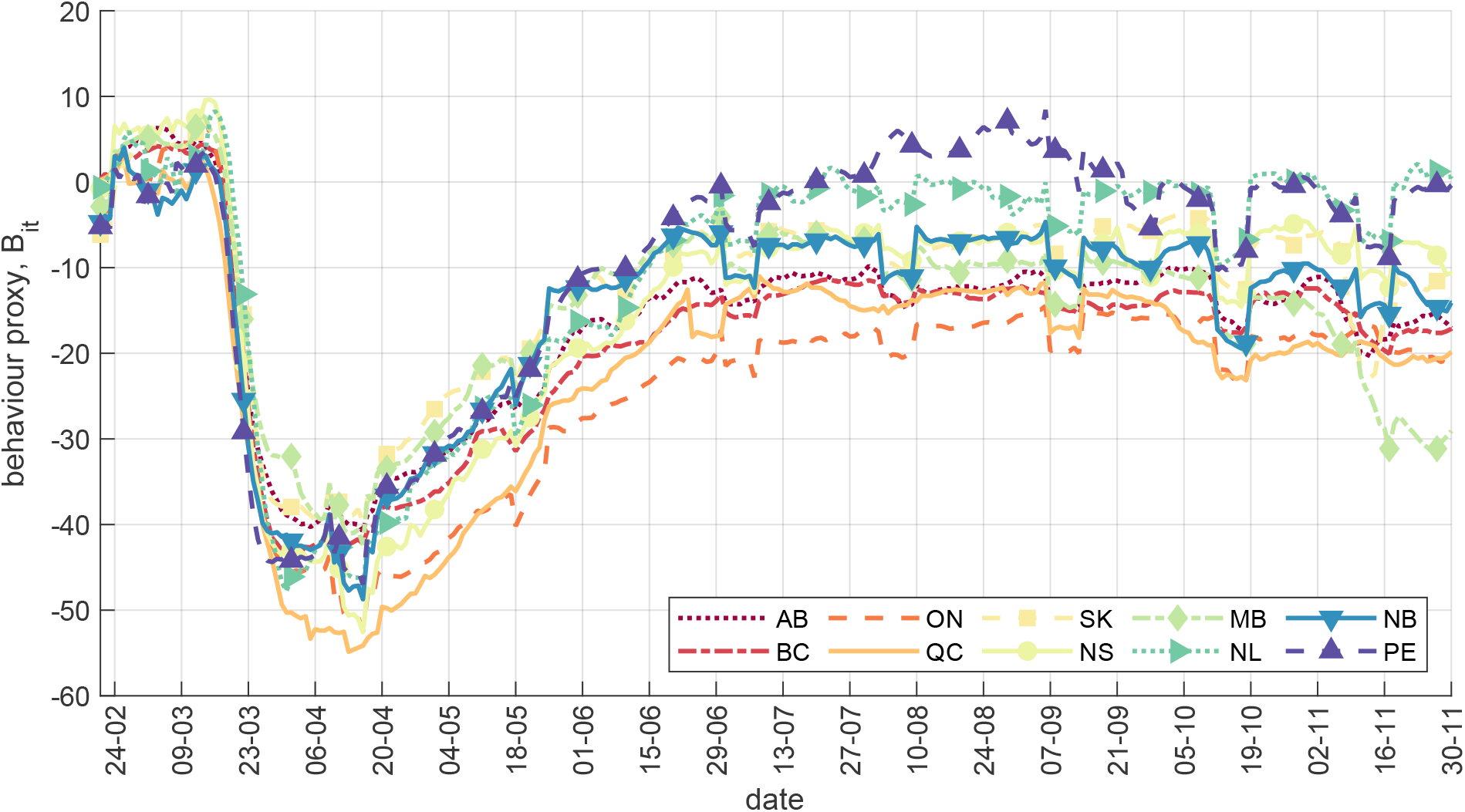
Canada – Behaviour proxy, *B*_*it*_ Notes: The Behaviour proxy *B*_*it*_ is the arithmetic average of the “retail”, “grocery and pharmacy”, and “workplaces” Google mobility indicators. Province-level 7-day moving averages are plotted.

**Figure C4:**
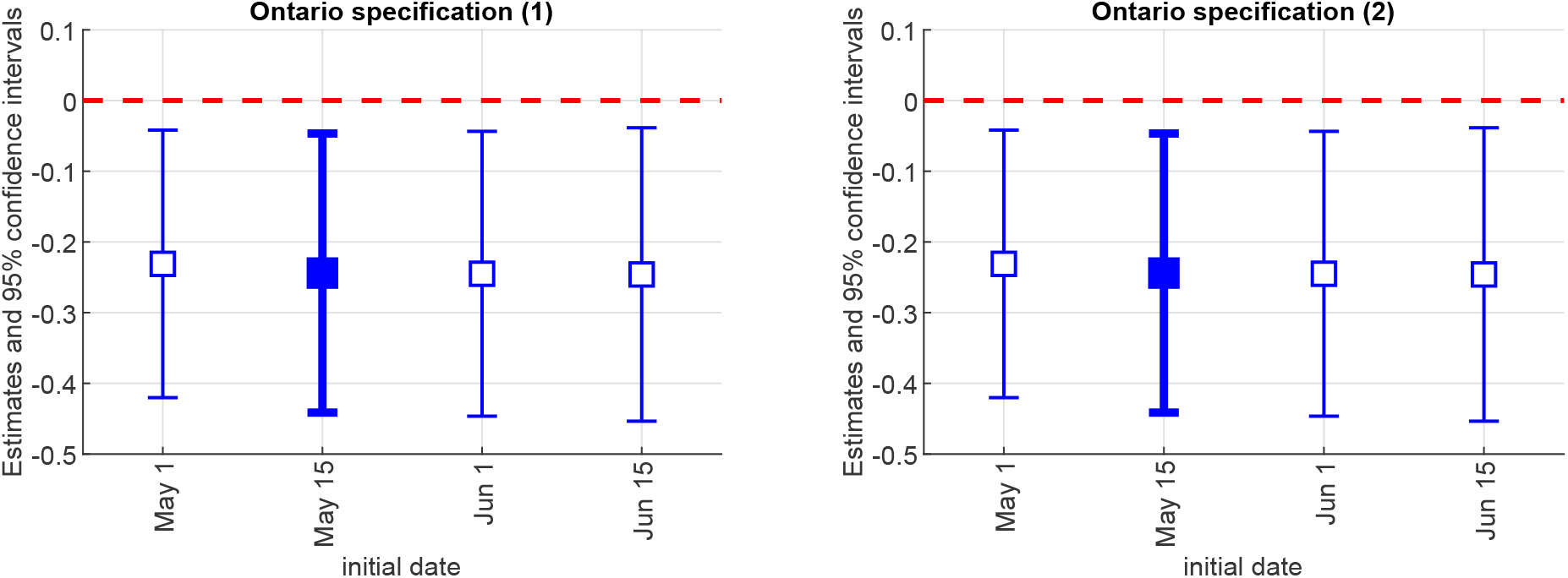
Ontario – different initial dates Notes: The figure plots the coefficient estimates on Mask 14, with 95% confidence intervals, from equation (1) for different initial dates of the sample. The baseline specifications (Table 1) use May 15. The left panel corresponds to column (1) in Table 1; the right panel corresponds to column (2) in Table 1.

**Figure C5:**
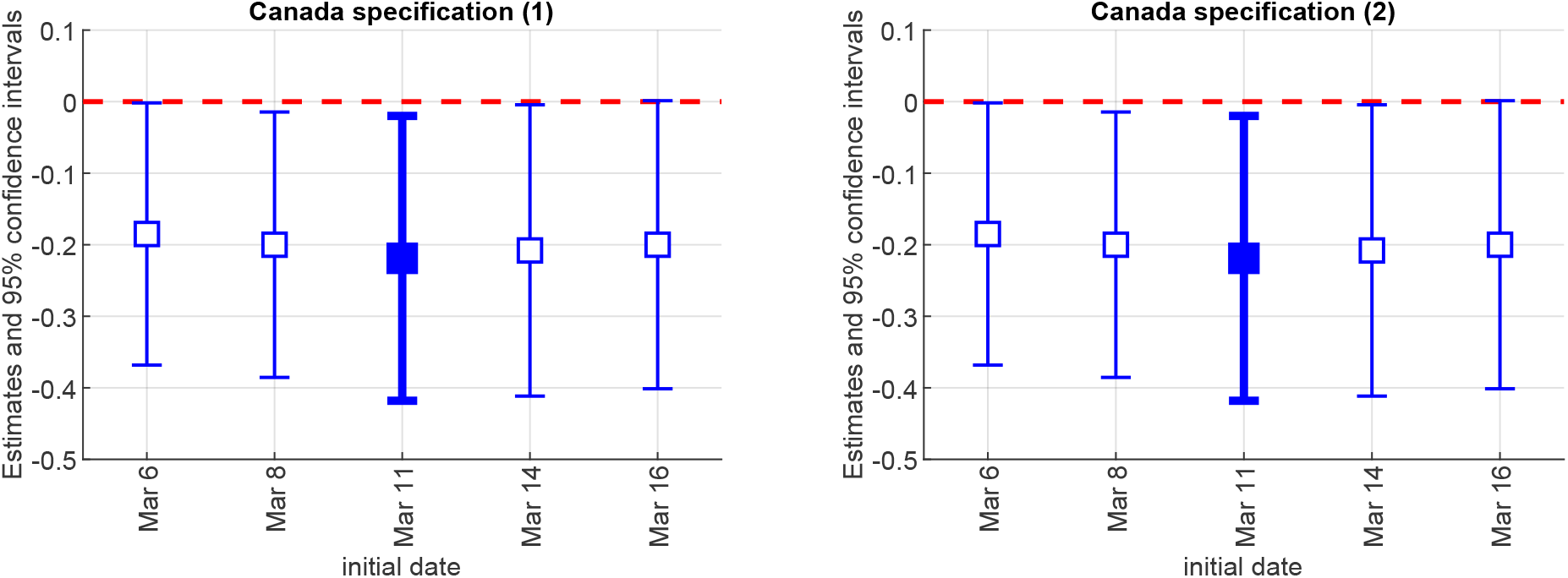
Canada – different initial dates Notes: The figure plots the coefficient estimates on Mask 14, with 95% confidence intervals, from equation (1) for different initial dates of the sample. The baseline specifications (Table 2) use March 11. The left panel corresponds to column (1) in Table 2; the right panel corresponds to column (2) in Table 2.

**Figure C6:**
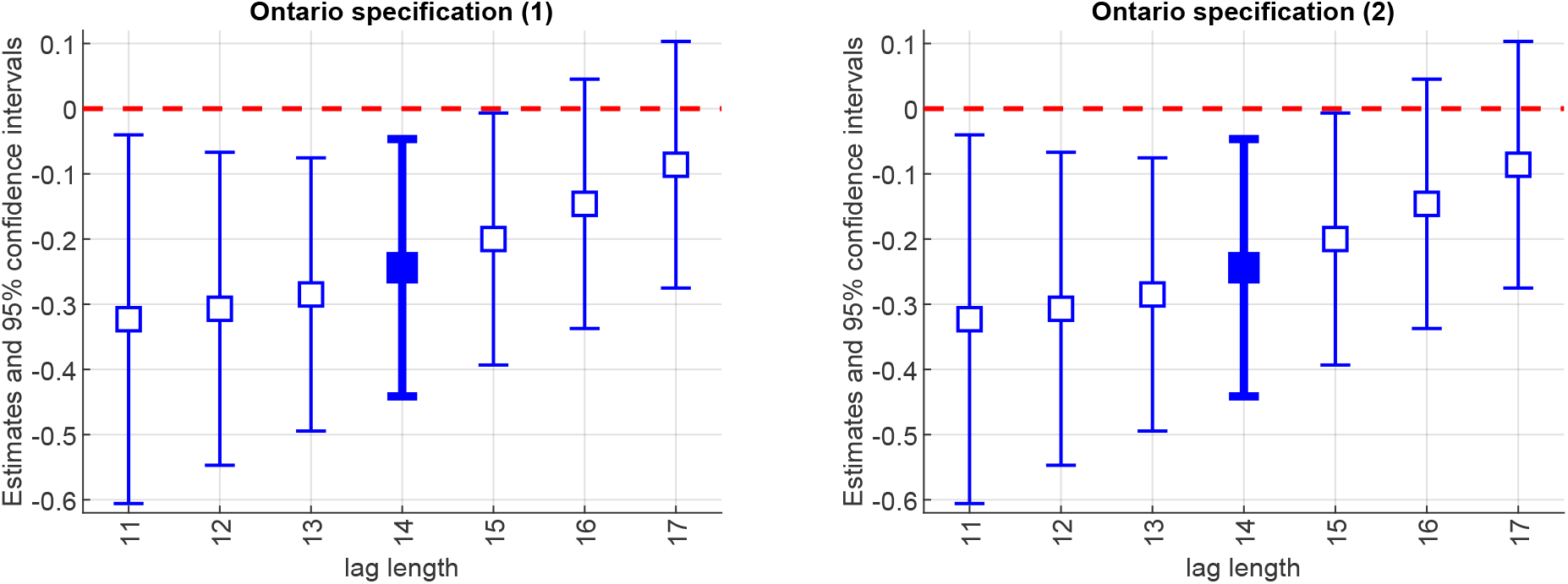
Ontario – different lag lengths Notes: The figure plots the coefficient estimates on Mask lag, with 95% confidence intervals, from equation (1) for different lag values. The baseline specifications (Table 1) use a lag of 14 days. The left panel corresponds to column (1) in Table 1; the right panel corresponds to column (2) in Table 1.

**Figure C7:**
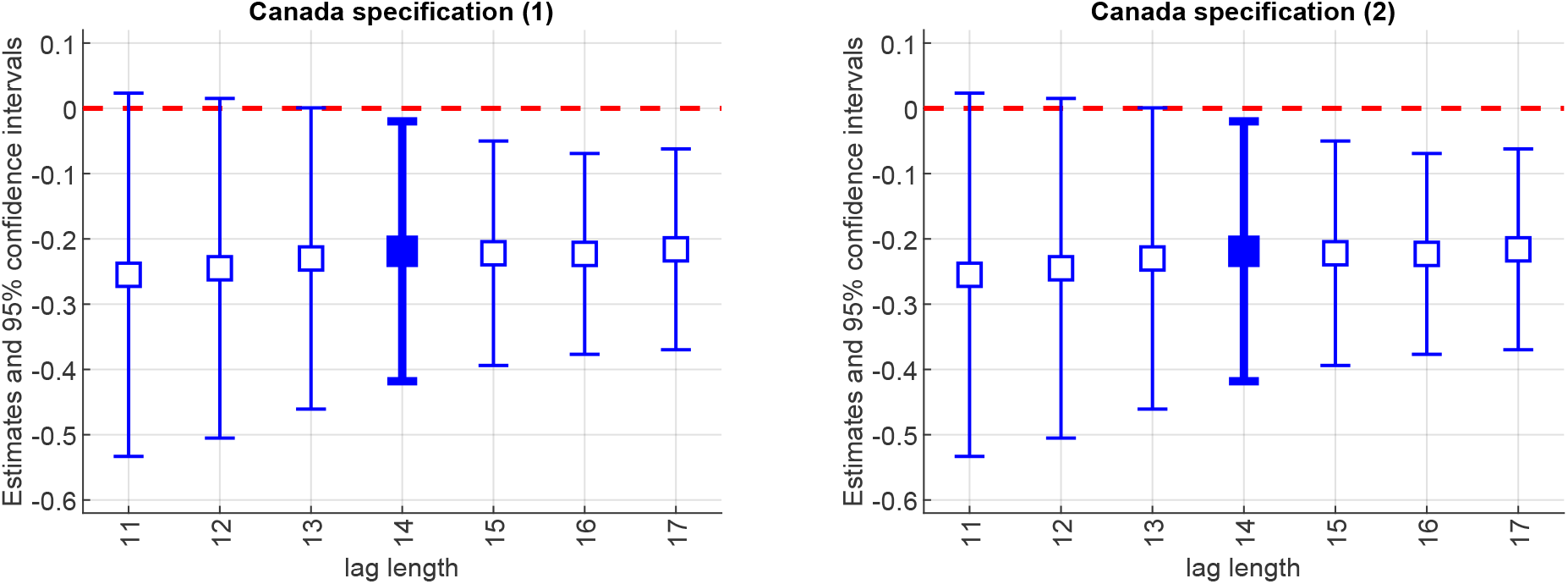
Canada – different lag lengths Notes: The figure plots the coefficient estimates on Mask lag, with 95% confidence intervals, from equation (1) for different lag values. The baseline specifications (Table 2) use a lag of 14 days. The left panels correspond to column (1) in Table 2; the right panels correspond to column (2) in Table 2.

**Figure C8:**
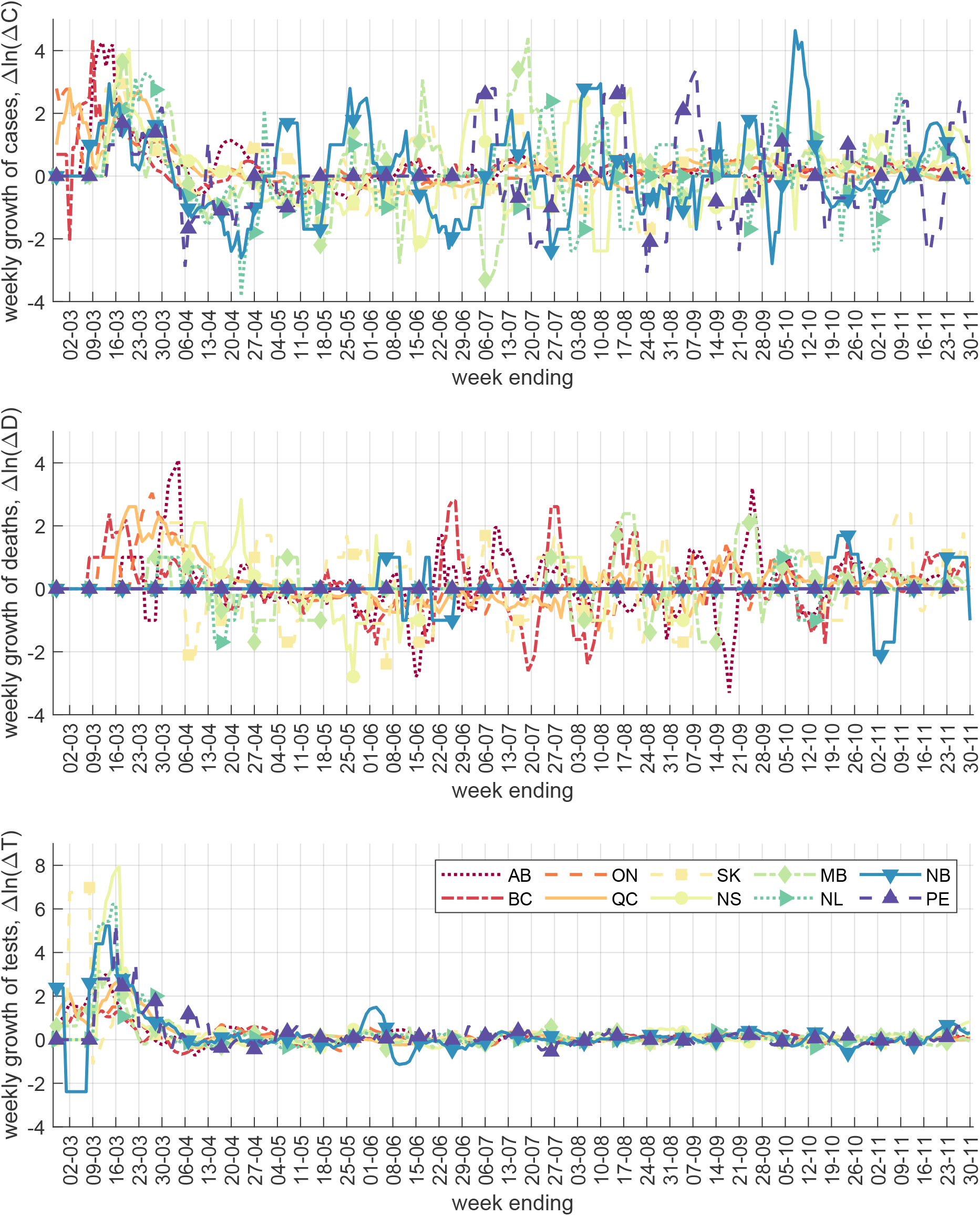
Canada – Weekly cases, deaths and tests (growth rate)

**Figure C9:**
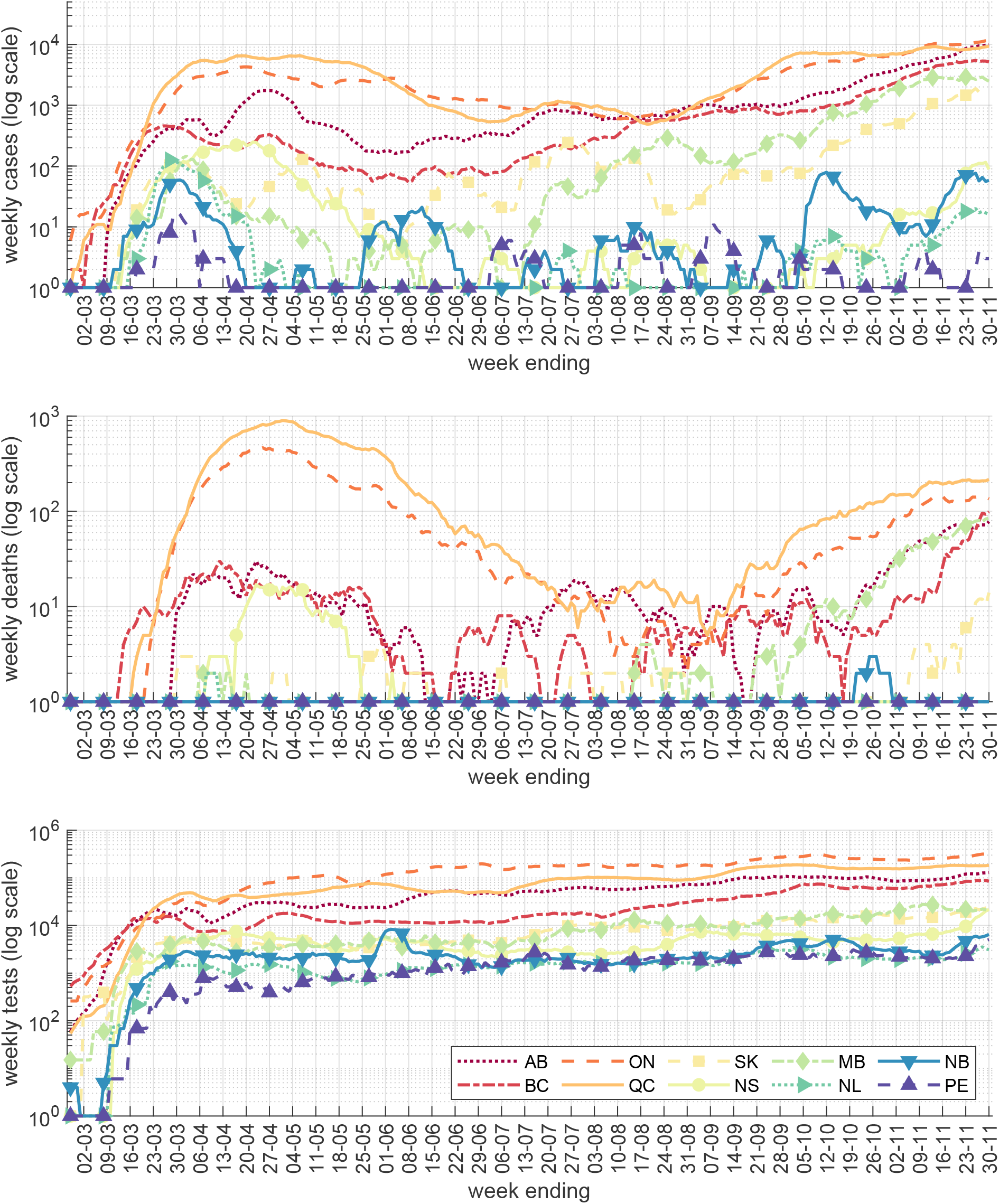
Canada – Weekly cases, deaths and tests (level)

**Figure C10:**
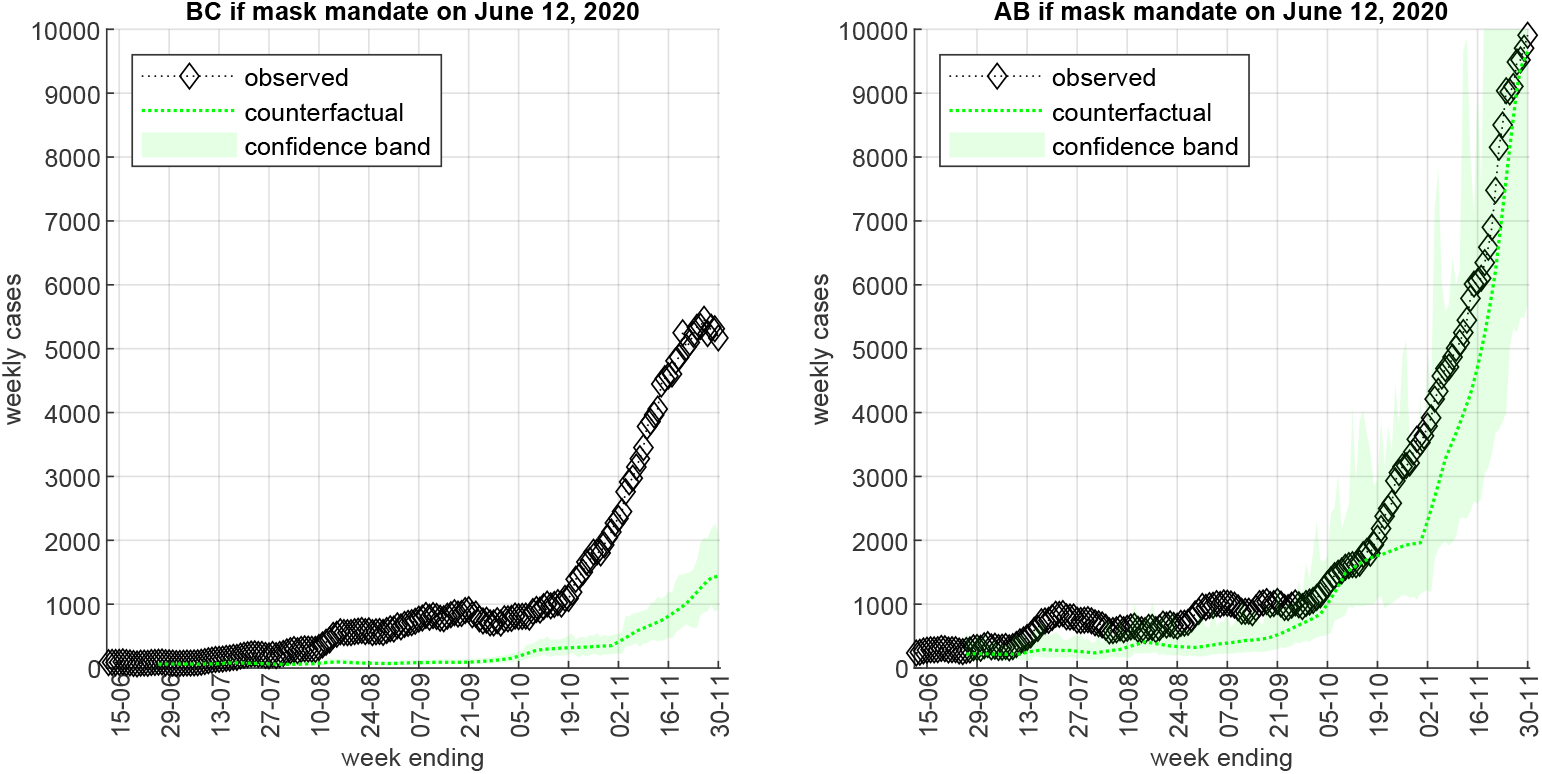
Counterfactuals – Earlier mask mandates in Alberta and British Columbia Notes: The figure assumes indoor mask mandate implementation on June 12 (date of the earliest mask mandate in Ontario) for the third and fourth largest provinces by population, specifically British Columbia (BC, left panel) and Alberta (AB, right panel). We use the estimates from column (2) of Table 2. The diamonds plot observed weekly cases from *t* −6 to *t*, the dotted lines plot the 7-day moving average of the counterfactual mean value, and the shaded areas are 5-95 percentile confidence bands.

**Figure C11:**
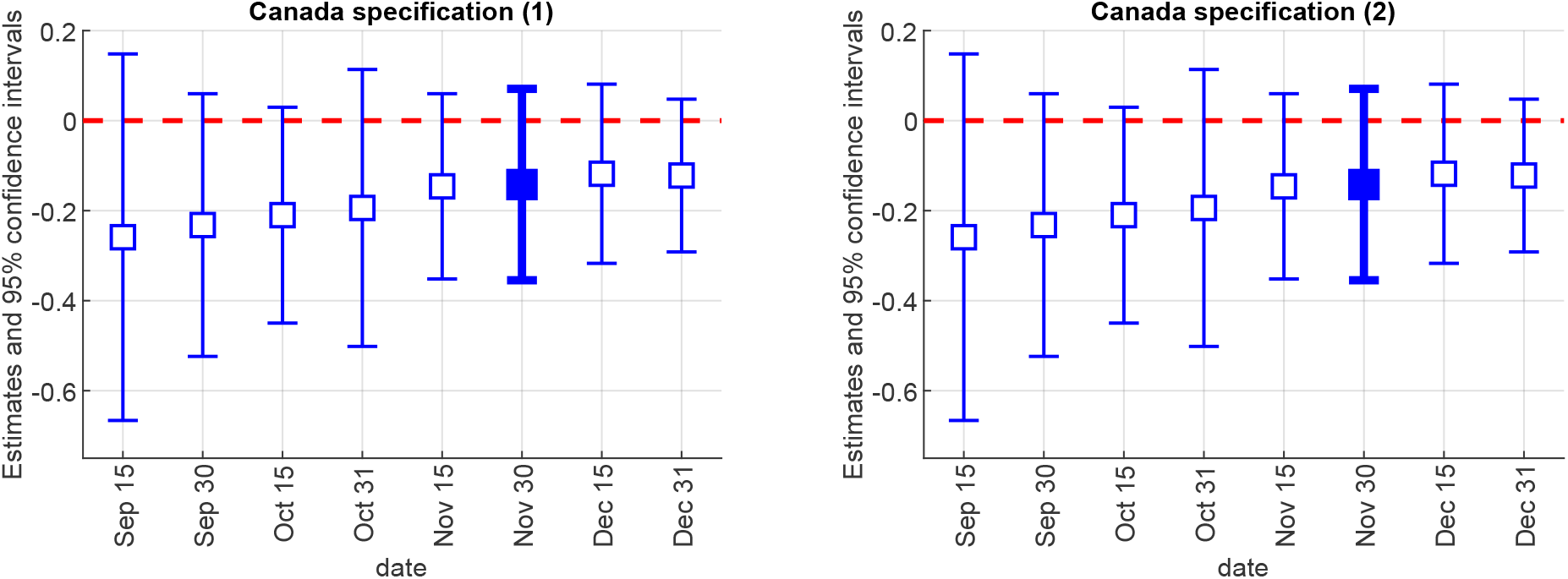
Canada – mask mandates and weekly deaths growth Notes: The figure plots the coefficient estimates on Mask 28, with 95% confidence intervals, from equation (1) using weekly deaths growth, Δlog(Δ*D*_*it*_), as the outcome variable, for different end dates of the sample. The baseline specifications (Table B17) use November 30. The left panel corresponds to column (1) in Table B17; the right panel corresponds to column (2) in Table B17.

## Appendix D. Definitions and Data Sources

**Table D1:**
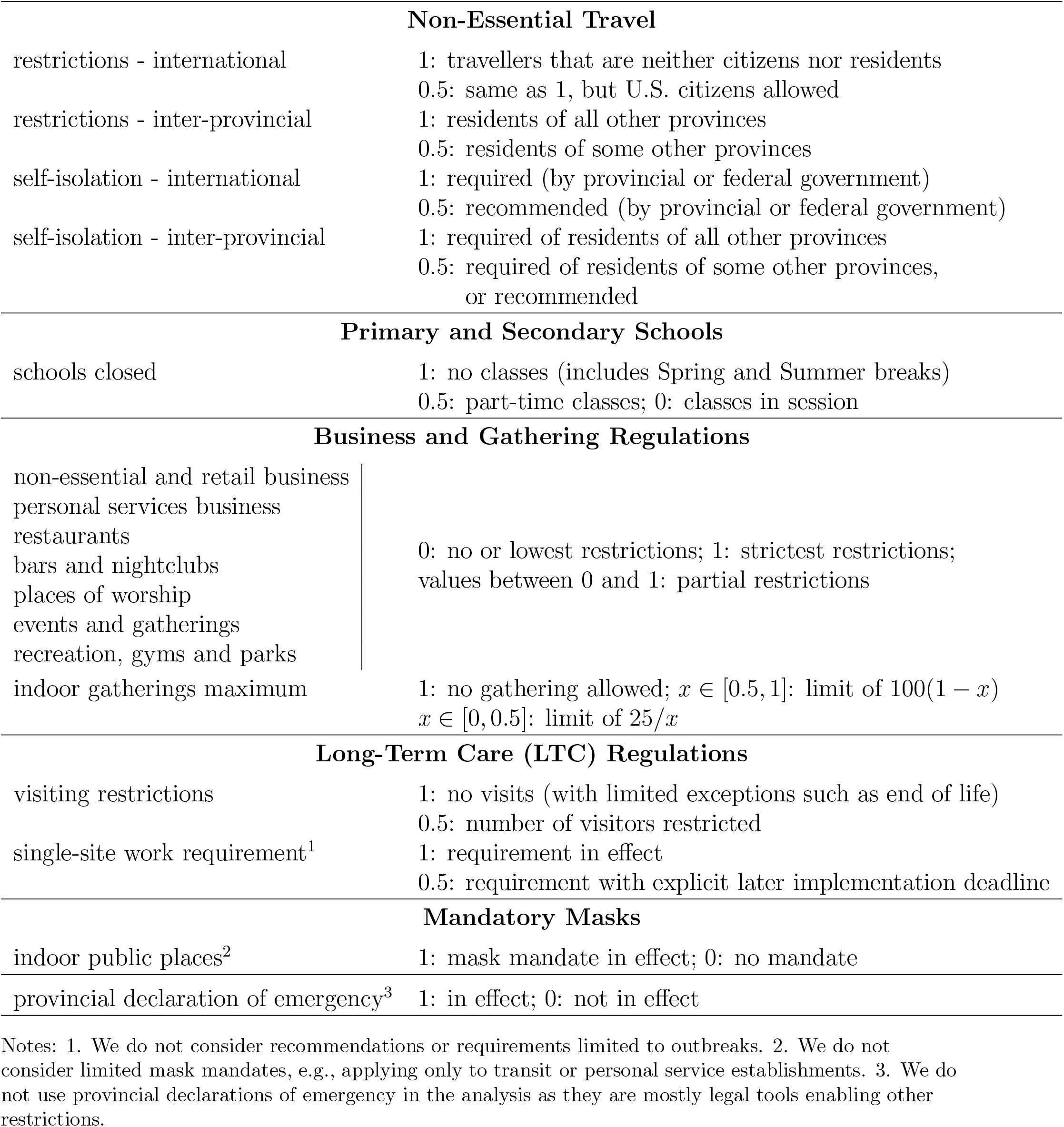
Policy indicators and aggregates

**Table D2:** Ontario public health regions – date of mask mandate

|  |  |  |
| --- | --- | --- |
| 1 | Algoma Public Health Unit | July 17, 2020 |
| 2 | Brant County Health Unit | July 20, 2020 |
| 3 | Chatham-Kent Health Unit | August 14, 2020 |
| 4 | Durham Region Health Department | July 10, 2020 |
| 5 | Eastern Ontario Health Unit | July 7, 2020 |
| 6 | Grey Bruce Health Unit | July 17, 2020 |
| 7 | Haldimand-Norfolk Health Unit | August 1, 2020 |
| 8 | Haliburton, Kawartha, Pine Ridge District Health Unit | July 13, 2020 |
| 9 | Halton Region Health Department | July 22, 2020 |
| 10 | Hamilton Public Health Services | July 20, 2020 |
| 11 | Hastings and Prince Edward Counties Health Unit | July 10, 2020 |
| 12 | Huron Perth District Health Unit | July 17, 2020 |
| 13 | Kingston, Frontenac and Lennox & Addington Public Health | June 27, 2020 |
| 14 | Lambton Public Health | July 31, 2020* |
| 15 | Leeds, Grenville and Lanark District Health Unit | July 7, 2020 |
| 16 | Middlesex-London Health Unit | July 18, 2020 |
| 17 | Niagara Region Public Health Department | July 31, 2020 |
| 18 | North Bay Parry Sound District Health Unit | July 24, 2020 |
| 19 | Northwestern Health Unit | August 17, 2020 |
| 20 | Ottawa Public Health | July 7, 2020 |
| 21 | Peel Public Health | July 10, 2020 |
| 22 | Peterborough Public Health | August 1, 2020 |
| 23 | Porcupine Health Unit | July 23, 2020 |
| 24 | Region of Waterloo, Public Health | July 13, 2020 |
| 25 | Renfrew County and District Health Unit | July 14, 2020 |
| 26 | Simcoe Muskoka District Health Unit | July 13, 2020 |
| 27 | Southwestern Public Health | July 31, 2020 |
| 28 | Sudbury & District Health Unit | July 17, 2020 |
| 29 | Thunder Bay District Health Unit | July 24, 2020 |
| 30 | Timiskaming Health Unit | July 24, 2020 |
| 31 | Toronto Public Health | July 7, 2020 |
| 32 | Wellington-Dufferin-Guelph Public Health | June 12, 2020 |
| 33 | Windsor-Essex County Health Unit | June 26, 2020 |
| 34 | York Region Public Health Services | July 17, 2020 |

\*Lambton Public Health did not enact a region-wide mask mandate until the provincial mandate on October 3. However, the City of Sarnia, with 58% of Lambton's population (2016 Census), enacted a mask mandate on July 31. The mask variable for Lambton is coded as 0.5 from July 31 to October 2, 2020.

**Table D3:** Canadian provinces – date of mask mandate

|  |  |  |
| --- | --- | --- |
| 1 | Alberta<sup>1</sup> | August 1, 2020 |
| 2 | British Columbia | November 19, 2020 |
| 3 | Manitoba<sup>2</sup> | September 28, 2020 |
| 4 | New Brunswick | October 9, 2020 |
| 5 | Newfoundland and Labrador | August 24, 2020 |
| 6 | Nova Scotia | July 31, 2020 |
| 7 | Ontario<sup>3</sup> | see Table D2 |
| 8 | Prince Edward Island | November 20, 2020 |
| 9 | Quebec | July 18, 2020 |
| 10 | Saskatchewan<sup>4</sup> | November 6, 2020 |

Notes: 1. Regional mandates: Calgary, Edmonton and surroundings (coded as 0.75), provincial mandate as of Dec. 8. 2. Regional mandates: Aug. 24–Sep. 17 (coded as 0.25), Sep. 28–Nov. 1 (coded as 0.5) and Nov. 2–Nov. 11 (coded as 0.75); provincial mandate as of Nov. 12. 3. Regional (PHU) mandates: see Table D2; provincial mandate as of Oct. 3. 4. Regional mandates: Nov. 6–Nov. 18 (coded as 0.75 and 0.95), provincial mandate as of Nov. 19.

**Table D4:** Canada COVID-19 official data sources

| Province | Cases | Deaths | Tests |
| --- | --- | --- | --- |
| Alberta (AB) | link | link | link |
| British Columbia (BC) | link | link | link |
| Ontario (ON) | link | link | link |
| Quebec (QC) | link | link | link |
| Saskatchewan (SK) | link<sup>1</sup> | link | link |
| Nova Scotia (NS) | link | link | link |
| Manitoba (MB) | link | link | link<sup>2</sup> |
| Newfoundland and Labrador (NL) | link<sup>1</sup> | link | link |
| New Brunswick (NB) | link | link | link |
| Prince Edward Island (PE) | link | link | link |

Notes: 1. Saskatchewan and Newfoundland and Labrador do not revise their posted data series. We made manual data adjustments based on subsequent revisions announced in government bulletins. 2. The Manitoba testing numbers were manually collected from the provincial government’s COVID-19 bulletins.

### Weather

We downloaded historical weather data for the largest city in each province from Weather Canada. The data provide daily information on 11 variables: maximum temperature (°C), minimum temperature (°C), average temperature (°C), heating degree-days, cooling degree-days, total rain (mm), total snow (cm), total precipitation (mm), snow on the ground (cm), direction of maximum wind gust (tens of degrees), and speed of maximum wind gust (km/h). We only use the temperature and precipitation data in Table B12 as possible factors determining outside vs. inside activity.

### News

We collected data from *ProQuest Canadian Newsstream*, a subscription service to all major and small-market daily or weekly Canadian news sources. We recorded the number of search results for each day from Feb. 1, 2020 to Nov. 30, 2020 by querying the database for the keywords “Coronavirus” or “COVID-19”. We only counted the results with source listed as “newspaper” since other sources such as blogs or podcasts tend to duplicate the same original content.

**Table D5:** YouGov Survey Questions

| Survey item | Question |
| --- | --- |
| i12_health_2 | Washed hands with soap and water |
| i12_health_3 | Used hand sanitizer |
| i12_health_6 | Avoided going out in general |
| i12_health_12 | Avoided small social gatherings (not more than 2 people) |
| i12_health_13 | Avoided medium-sized social gatherings (between 3 and 10 people) |
| i12_health_14 | Avoided large-sized social gatherings (more than 10 people) |
| i12_health_15 | Avoided crowded areas |
| i12_health_20 | Avoided touching objects in public (e.g., elevator buttons or doors) |

Notes: The data source is YouGov. Possible responses to each survey item are “Always”, “Frequently”, “Sometimes”, “Rarely”, and “Not at all”. For Table B14, we create a binary variable taking value 1 if the response is “Always” and 0 otherwise. For Table B15, we create a binary variable taking value of 1 if the respondent answered either “Always” or “Frequently” and 0 otherwise.

All data used in the paper are available at github.com/C19-SFU-Econ/data.

## Appendix E. Lags Determination

As discussed in Section 3.1, we assume a lag of 14 days between a change in policy or behaviour and its hypothesized effect on weekly case growth, and lag of 28 days between such a change and its effect on weekly death growth.

First, we consider the lag between infection and a case being reported. As most identified cases of COVID-19 in Canada are symptomatic, we focus on symptomatic individuals. For most provinces cases are listed according to the date of report to public health. In provinces where the dates instead refer to the public announcement, we shifted them back by one day, as announcements typically contain the cases reported to public health on the previous day. The relevant lag therefore has two components:

1. Incubation period: most studies suggest an average incubation period of 5-6 days (e.g. 5.2 days in Li et al. (2020), 5.5 days in Lauer et al. (2020), 5.6 days in Linton et al. (2020), 6.4 days in Backer et al. (2020)).
2. Time between symptoms onset and reporting the case to public health: the Ontario data contain an estimate of the symptom onset date (“episode date”) for each case. For our sample period, the average difference between the date of report and the episode date is 4.7 days, including only values from 1 to 28 days. We assume that the lags in Ontario and in other provinces are similar, and use a value of 5 days between symptom onset and report to public health authorities.

Altogether, this implies that the average lag between infection and the reporting of a positive case to public health is approximately 10–11 days. Since awareness of a policy is often partial on its first day in effect, we use a lag of 11 days. Second, we consider the effect of weekly averaging on the appropriate lag for our analysis. Suppose a policy or behavioural change starts on date *t*, impacting the daily growth in infections between dates *t* − 1 and *t* and in each subsequent day. Then, assuming a lag of 11 days between infection and case reporting, case counts *C* are affected from date *t* + 11 onward. Our outcome variable Δ log(ΔC) thus would react to the original policy or behavioural change on date *t* + 11. The change is complete on *t* + 23, when the week from *t* + 17 to *t* + 23 is compared to the week from *t* + 10 to *t* + 16. The midpoint of the change is *t* + 17. Choosing a lag of *l* days implies that the policy/behaviour variable phases in from *t* + *l* to *t* + *l* + 6.

To match the midpoint of this phase-in to the midpoint of the change in the outcome variable, we set *l* = 14. The chosen lag also matches the lag used by other authors who study COVID-19 policy interventions, e.g., Chernozhukov et al. (2021). We explore the sensitivity of our results to different lag lengths in Section 4.4.

Regarding deaths, our data are, in most cases, backdated (revised by the authorities expost) to the actual date of death. The medical literature suggests that the mean time from symptom onset to death is around 19 days (20 days in Wu et al., 2020; 17.8 days in Verity et al., 2020; 20.2 days when accounting for right truncation in Linton et al., 2020; 16.1 days in Sanche et al., 2020), that is, two weeks longer than our estimate of the time from symptom onset to reporting of a positive test result. We accordingly set the lag used in our analysis of the death growth rate in Appendix A to 28 days.

## Footnotes

1 Hatzius et al. (2020) estimate that a national mask mandate in the U.S. could replace alternative restrictions costing 5% of GDP.

2 In Canada, the Chief Public Health Officer (CPHO) shifted from not recommending mask use to describing it as a measure that asymptomatic individuals “can take to protect others” on Apr. 6, 2020, and only officially recommended mask wearing on May 20, 2020. However, even after the official CPHO recommendation, messaging remained mixed: for example, in defending British Columbia’s lack of mask mandate, the Provincial Health Officer described masks as the “least effective layer of protection” as late as on July 23 and Aug. 10, 2020.

3 We show mask usage for the U.S. and Germany because of related work on mask mandates by Chernozhukov et al. (2021) and Mitze et al. (2020). We show Australia as an example of a country that did not mandate masks (except in Melbourne). See Hatzius et al. (2020) for more cross-country comparisons.

4 The Ontario Ministry of Health and Long-Term Care (2021) describes a public health unit as “an official health agency established by a group of urban and rural municipalities to provide a more efficient community health program, carried out by full-time, specially qualified staff.”

5 Chernozhukov et al. (2021) use U.S. state-level variation in the timing of mask mandates for employees in public-facing businesses and find that these mandates are associated with a 10 percentage point reduction in the weekly growth rate of cases. Mitze et al. (2020) use a synthetic control approach and compare the city of Jena and six regions in Germany that adopted a face mask policy before their respective state mandate. The authors find that mandatory masks reduced the daily growth rate of cases by about 40%.

6 See also Abaluck et al. (2020) who compare countries with pre-existing ‘wear mask when sick’ norms and countries without such norms and report 8 percentage points lower average daily case growth rate in the former countries (S. Korea, Japan, Hong Kong and Taiwan).

7 The provinces differ in the ease of accessibility of their official COVID-19 data time series. In some cases, we located and used the hidden json sources feeding the public dashboard charts. In a few instances in which data were not available, we used the numbers reported in the daily provincial government announcements. All data sources are referenced and web-linked in Appendix Table D4.

8 Additional survey data on mask usage is described and used in Section 4.3.

9 Decisions about the former were made at the PHU level, while decisions about the latter were made by the province, which classified PHUs into groups.

10 The reports are available at www.google.com/covid19/mobility/.

11 Each of these divisions is either entirely (in most cases) or predominantly located within a single public health unit (PHU). In the cases where a PHU consists of multiple divisions, 2016 Census population numbers were used as weights to compute the PHU geo-location behaviour index.

12 We also report results using the growth rate of deaths as supplementary analysis in Appendix A, with the death outcome variable defined analogously.

13 To handle zero weekly values, which mostly occur in the smaller regions, we replace log(0) with −1 as in Chernozhukov et al. (2021). We also check the robustness of our results by adding 1 to all Δ*C*_*it*_ before taking logs, replacing log(0) with 0, or using population-weighted least squares; see Tables B6 and B9.

14 There was no PHU-wide mask mandate in Lambton Health until Ontario’s province-wide mandate on Oct. 3. However, Lambton’s main city Sarnia enacted a mask mandate on July 31.

15 Policies and cases in a region may also impact case growth in neighbouring regions. Because of data limitations, we are unable to directly address this concern in the current paper. However, we note that this issue should tend to attenuate the estimated effect of policies if a fraction of cases are unaffected by policy changes in the jurisdiction where they are counted because of infections occurring elsewhere. Moreover, measurement error may be introduced if policy changes elsewhere affect a jurisdiction’s cases. Both these effects would work against finding statistically significant results.

16 The daily numerical values for each of the basic policy indicators and the 5 policy aggregates for each province and date are available on the project’s Github repository.

17 Alternative ways of computing the standard errors are explored in Section 4.4.

18 We drop the “transit”, “parks”, and “residential” location indicators because, respectively, 12.9%, 17.5%, and 2% of the observations are missing in the provincial data, and 26%, 56%, and 8.4% are missing in the Ontario data. Furthermore, the “transit” and “residential” variables are highly correlated with the three indicators included in our behaviour proxy *B*_*it*_, and the “parks” indicator does not have clear implication for COVID-19 outcomes.

19 In the Ontario analysis, 1% of the *B*_*it*_ values were imputed via linear interpolation.

20 The correlation between weekly tests Δ*T*_*it*_ and the presence of a mask mandate is low in Ontario and moderately positive across provinces (see Tables B3 and B4); it is present because of the coincident expansion of mask mandates across regions and gradually increasing testing rates over time. To the extent that unobserved changes in testing strategy may affect our estimates, we believe that they would work against our findings of NPI effects, as restrictions may coincide with increased testing. However, any such effect is likely minor: one-time changes in testing only temporarily affect the growth rate of detected new cases. Moreover, for our main Ontario analysis, provincial guidelines limit PHUs’ discretion in testing effort.

21 Mask mandates, regulations on businesses and gatherings and the growth rate of tests vary at the PHU level. The long-term care and school policies are at the provincial level. Travel policies do not vary in the sample period and hence are omitted from the analysis with Ontario PHU data.

22 Table B7 in the Appendix reports alternative standard error specifications: regular clustering at the PHU level (Stata command “cluster”), wild bootstrap standard errors clustered at the PHU level, and wild bootstrap standard errors two-way clustered by PHU and date.

23 In all tables, *Variable*_14 denotes the 14-day lag of *Variable*.

24 Using equations (1) and (3), a coefficient estimate 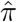 on Mask_14, as the latter changes from 0 to 1, corresponds to a exp(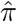) − 1 percent change in the ratio of current-week to past-week cases, Δ*C*_*it*_/Δ*C*_*it*−7_. For example, suppose weekly cases in weeks 1, 2 and 3 are 100, 150 and 225 respectively, showing a 50% weekly case growth. Our estimates suggest that, all else equal, a mask mandate implemented at the beginning of week 1 would reduce week 3 cases to 176 (instead of 225 in absence of mandate), with continued reductions in the next weeks.

25 School closures (School_14) have a negative coefficient. However, the estimates are only meaningfully different from zero in the specifications without time trend in Table B5. As seen on Fig. 2, school closures occurred in a very short period in March 2020, so we lack statistical power to separately identify their effect from that of other NPIs (e.g., travel restrictions) and the week fixed effects.

26 The YouGov data is available at https://yougov.co.uk/covid-19.

27 We use July 7, the mask mandate adoption date in Toronto and Ottawa, as mandate date for Ontario.

28 Since mask usage is reported only for specific dates within each survey wave, we use the Mask variable daily values for these same dates instead of the weekly moving average.

29 We also present results without time trend or including cubic time trend in days in Table B13.

30 The finding that the increase in mask usage among the “Always” respondents is larger than among the “Always” or “Frequent” respondents is consistent with some people switching from wearing masks “frequently” to “always”.

31 Hatzius et al. (2020) document that state mask mandates in the USA increased mask usage roughly by 25 percentage points in 30 days. The compliance with mask mandates may differ across countries or regions based on social norms, peer effects, political factors or the consequences of noncompliance.

32 If we take the increase of 27 percentage points in mask usage at face value, the full effect of mask wearing (treatment-on-the-treated) would be roughly triple our estimates. It could be larger still if there is desirability bias in answering the mask usage question, so that the actual increase in usage may be smaller than our estimate.

33 Seres et al. (2020) found that wearing masks increased physical distancing in a randomized field experiment at waiting lines outside German stores.

34 887 out of the 6,800 observations (13%) in Table 1 and 329 out of the 2,650 observations (12%) in Table 2 are affected, mostly in the smaller provinces or PHUs. When both Δ*C*_*it*_ and Δ*C*_*it*−7_ are zero, the weekly case growth rate is *Y*_*it*_ = 0.

35 Aggregating the 17 basic policy indicators into five groups mitigates this issue. Here, we test whether any remaining collinearity poses a problem.

36 Vancouver, BC; Calgary, AB; Saskatoon, SK; Winnipeg, MB; Toronto, ON; Montreal, QC; Moncton, NB; Halifax, NS; Charlottetown, PE; and St. John’s, NL.

37 de Chaisemartin and D’Haultfoeuille (2018), Goodman-Bacon (2018), Callaway and Sant’Anna (2021).

38 We use the Stata command *did_multiplegt* provided by the authors. The estimator is a weighted average, across time periods *t* and treatment values *d*, of difference-in-difference estimators comparing the outcome change observed in groups whose treatment status changes from *d* to some other value from *t* − 1 to *t*, to that observed in groups whose treatment status is *d* at both dates.

39 The standard error is large as the estimator relies on only 231 ‘switchers’ (N=1,162) unlike our full sample with N= 6,800.

40 All details and complete regression tables are available upon request.

